# Does Ventricular Substrate Play a Role in Incident Stroke? The Atherosclerosis Risk in Communities (ARIC) Study

**DOI:** 10.1101/2020.05.04.20090910

**Authors:** John A. Johnson, Kazi T. Haq, Katherine J. Lutz, Kyle K. Peters, Kevin A. Paternostro, Natalie E. Craig, Nathan Stencel, Lila Hawkinson, Maedeh Khayyat-Kholghi, Larisa G. Tereshchenko

**Author notes:** <u>Correspondence</u>: Larisa Tereshchenko, 3181 SW Sam Jackson Park Rd; UHN62; Portland, OR, 97239.

## Abstract

**Background:** —The goal of the study was to determine an association of ventricular substrate with thrombotic, cardioembolic, and hemorrhagic stroke.

**Methods:** —Participants from the Atherosclerosis Risk in Communities study with analyzable ECGs and no history of stroke were included (n=14,479; age 54±6 y; 55% female; 24% black). Ventricular substrate was characterized by cardiac memory [spatial QRS-T angle (QRS-Ta), sum absolute QRST integral (SAIQRST), spatial ventricular gradient magnitude (SVGmag)], premature ventricular contractions (PVC) and tachycardia-dependent intermittent bundle branch block (TD-IBBB) on 12-lead ECG recorded at visits 1-5. Incident strokes were adjudicated by physician reviewers. Cox proportional hazard risk models were constructed.

**Results:** —Over a median 24.5 y follow-up, there were 899 thrombotic, 400 cardioembolic, and 187 hemorrhagic strokes. After adjustment for cardiovascular disease (CVD) and its risk factors, atrial fibrillation / atrial substrate, and ECG-left ventricular hypertrophy, PVC (HR 1.72; 95%CI 1.02-2.92), QRS-Ta (HR 1.15; 95%CI 1.03-1.28), SAIQRST (HR 1.20; 95%CI 1.07-1.34) and time-updated SVGmag (HR 1.19; 95%CI 1.08-1.32) associated with cardioembolic stroke. After adjustment for CVD and its risk factors, PVC (HR 1.53; 95%CI 1.03-2.26), QRS-Ta (HR 1.08; 95%CI 1.01-1.16), SAIQRST (HR 1.07; 95%CI 1.01-1.14), and time-updated SVGmag (HR 1.11; 95%CI 1.04-1.19) associated with thrombotic stroke. In fully adjusted time-updated Cox model QRS-Ta (HR 1.20; 95%CI 1.04-1.38), SAI QRST (HR 1.23; 95%CI 1.06-1.43), SVGmag (HR 1.23; 95%CI 1.06-1.43) associated with hemorrhagic stroke, whereas TD-IBBB trended (HR 1.84; 95%CI 0.25-13.33).

**Conclusions:** —PVC burden reflected by cardiac memory is associated with ischemic stroke. Transient cardiac memory (likely through TD-IBBB) precedes hemorrhagic stroke.

## Introduction

Stroke remains the leading cause of long-term disability and the 5^th^ leading cause of death in the United States.^1^ Despite declining stroke incidence in adults over 65 years of age^2^, the global lifetime risk of stroke increased from 22.8% in 1990 to 24.9% in 2016.^3^ Large portions, approximately 90.5% of stroke risk, could be attributed to traditional cardiovascular risk factors, such as hypertension, obesity, hyperlipidemia, hyperglycemia, renal dysfunction, smoking, and sedentary lifestyle, as well as air pollution.^4^ Nevertheless, a recent genome-wide association study of stroke identified 22 previously unknown genetic loci, pointing towards additional, currently unrecognized mechanisms of stroke.^5^ The most accurate stroke risk prediction is provided by CHA_2_DS_2_-VASc and P_2_-CHA_2_DS_2_VASc risk scores^6^, in persons with and without^7^ atrial fibrillation (AF). However, C-statistics for the stroke risk scores remain suboptimal (~0.6 to 0.7). Thus, it is essential to understand additional factors beyond those already accounted for in the risk assessment models.

Several studies reported an association of both premature ventricular complexes (PVCs)^8, 9^, and left ventricular hypertrophy (LVH)^10^, with incident stroke. This suggests an abnormal ventricular substrate, with or without atrial myopathy, may serve as an independent risk factor. Furthermore, it was previously shown that left ventricular interstitial fibrosis is associated with interatrial conduction abnormalities, characterized by P-wave indices.^11^ However, at present, the role of a ventricular substrate in stroke mechanisms is not entirely clear.

Global electrical heterogeneity (GEH) recently emerged as a global measure of an abnormal electrophysiological substrate in the ventricles of the heart.^12^ GEH is a concept based on Wilson’s spatial ventricular gradient (SVG) theory.^13^ It was mathematically demonstrated that the sum of the QRS and T vectors should theoretically be zero if depolarization and repolarization occurred in the same manner. By measuring the SVG, one could quantify the global dispersion of activation and recovery. SVG is independent of the type of ventricular activation.^14, 15^ Therefore, it can be measured in individuals with or without AF or ventricular conduction abnormalities, enabling a broad study of ventricular substrates. An association of GEH with stroke has not been previously studied.

We conducted this study with the goal of uncovering novel mechanisms of stroke, to determine an association of ventricular substrate (quantified by GEH and traditional ECG metrics) with thrombotic, cardioembolic, or hemorrhagic stroke subtypes. We hypothesized that GEH is associated with incident stroke in the Atherosclerosis Risk in Communities (ARIC) study participants.

## Methods

The ARIC Study data are available through the National Heart, Lung, and Blood Institute’s Biological Specimen and Data Repository Information Coordinating Center (BioLINCC)^16^, the National Center of Biotechnology Information’s database of Genotypes and Phenotypes (dbGaP)^17^, and via ARIC Coordinating Center at the University of North Carolina—Chapel Hill^18^. Informed consent was obtained from all study participants prior to enrollment. The study was approved by the Oregon Health & Science University Institutional Review Board.

### Study population

The ARIC study enrolled 15,792 participants (age 45-64 y) between 1987 and 1989.^19^ In this study, we included ARIC study participants with recorded resting 12-lead ECG and measured global electrical heterogeneity (GEH)^12^; n=15,776. ECGs were recorded during visits 1-5. Visit 1 was coordinated in 1987-1989, visit 2 in 1990-1992, visit 3 in 1993-1995, visit 4 in 1996-1998, and visit 5 in 2011-2013. We excluded participants with a prevalent stroke (defined^20^ as previous stroke or transient ischemic attack identified by a standardized interview) that occurred before the 1^st^ ECG GEH measurement (n=278). We further excluded those who were not white or black from all study sites, and nonwhite from Washington County and Minneapolis field centers (due to the relatively small sample size, n=101). Finally, we excluded participants with missing covariates or outcome (n=918). The final sample included 14,479 participants (Figure 1).

**Figure 1.**
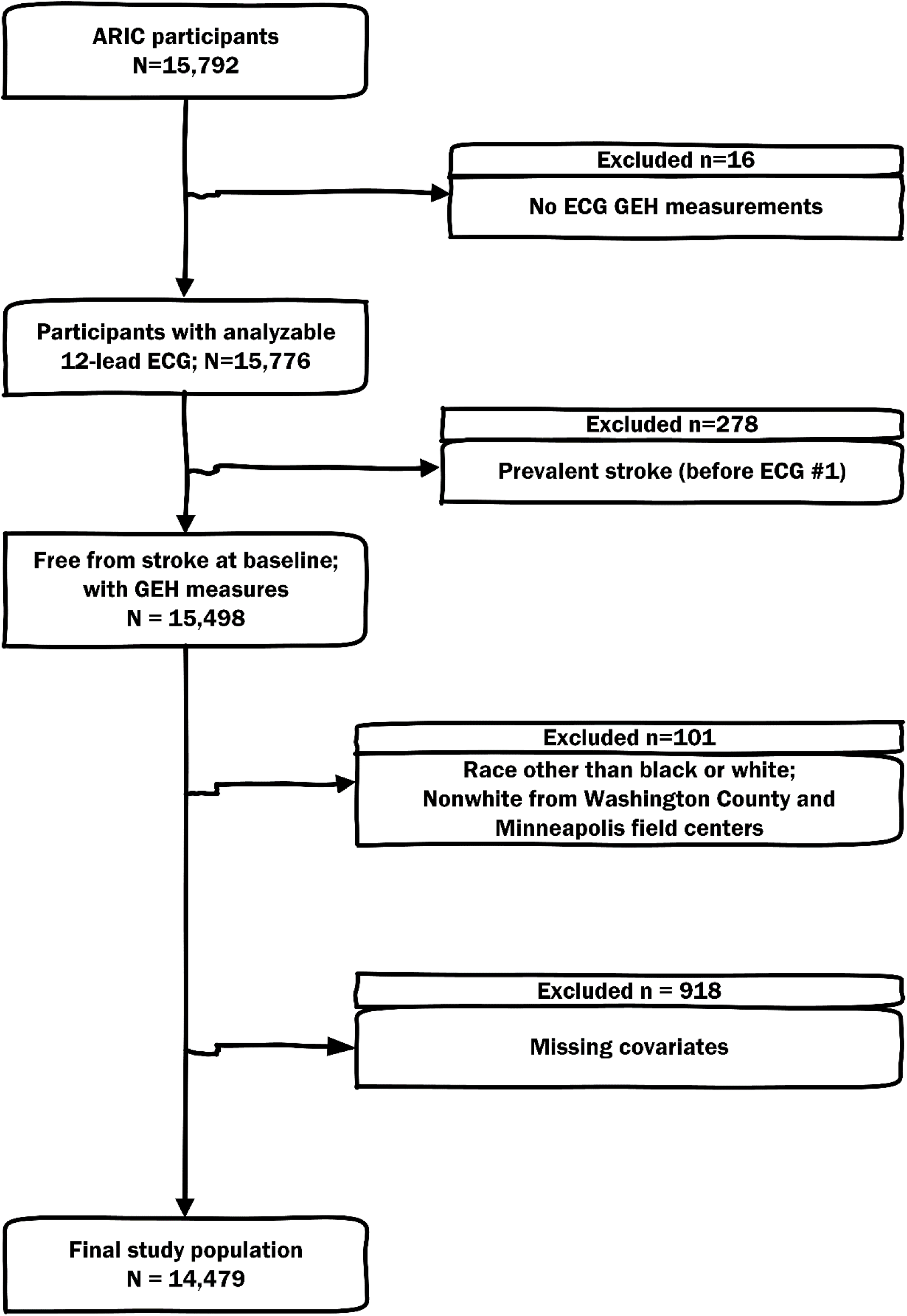
Flowchart of study cohort development.

### Exposure: Global Electrical Heterogeneity and other measures of ventricular substrate

We analyzed resting 12-lead ECGs of the first five study visits.^12, 21-23^ Ventricular substrate was characterized by novel ECG measures (GEH), by the presence of premature ventricular complexes (PVCs) on 10-second 12-lead ECG, and by traditional ECG measures [QRS duration, Bazett-corrected QTc interval, electrocardiographic left ventricular hypertrophy (ECG-LVH), bundle branch block (BBB) or interventricular conduction delay (IVCD)].

Each beat on a 10-second 12-lead ECG was manually labeled by at least two physicians or physicians-in-training investigators (JJ, KJL, KP, KAP, NC, LH, MKK, LGT) at the Oregon Health & Science University (Tereshchenko laboratory)^21-23^, and PVCs were identified. We created a time-coherent median beat with identified isoelectric heart vector origin point.^24^ A median beat was comprised of only one (dominant) type. In this study, we included three groups of median beats. Normal (N) group included median beats conducting from atria to ventricles (normal sinus, atrial paced, junctional, and ectopic atrial median beats). The ventricular (V) group included median beats with ventricular activation originating in ventricles (ventricular paced, and both atrial and ventricular paced median beats). The supraventricular (S) group included median beats of AF or atrial flutter with an atria-to-ventricle type of ventricular conduction.

GEH was measured by spatial QRS-T angle, SAI QRST, and SVG magnitude, azimuth, and elevation.^25^ We used two approaches for measurement of SVG vectors and QRS-T angles: area-based, and peak-based.^21, 24, 25^ The open-source MATLAB (MathWorks, Natick, MA, USA) software code for GEH and the heart vector origin measurement is provided at https://physionet.org/physiotools/geh and https://github.com/Tereshchenkolab/Origin. We used previously selected sex- and race-stratified thresholds for abnormal GEH measurements^12^, to comprise the GEH risk score ranging from 0 to 5 abnormal GEH metrics.

QRS duration, QTc, R_aVL_, and S_V3_ amplitudes were measured by the 12 SL algorithm as implemented in the Magellan ECG Research Workstation V2 (GE Marquette Electronics, Milwaukee, WI). Cornell voltage was calculated as the sum of R_aVL_ and S_V3_ amplitudes. ECG-LVH was defined as a sex-adjusted Cornell product (Cornell voltage times QRS duration)^26^ >2440 mm*ms.

BBB was identified by the Minnesota Code^27^ (left bundle branch block code 7.1, right bundle branch block code 7.2 and 7.3, fascicular block code 7.6-7.7, bi-fascicular block code 7.8). IVCD was defined as a QRS duration above 120 ms in the absence of BBB criteria.

Tachycardia-dependent intermittent BBB (TD-IBBB) was diagnosed on a 12-lead ECG if an intermittent BBB morphology appeared on a normal sinus beat with shortened RR’ interval or PAC with aberrant ventricular conduction (Supplemental Figures 1 and 2).

**Figure 2.**
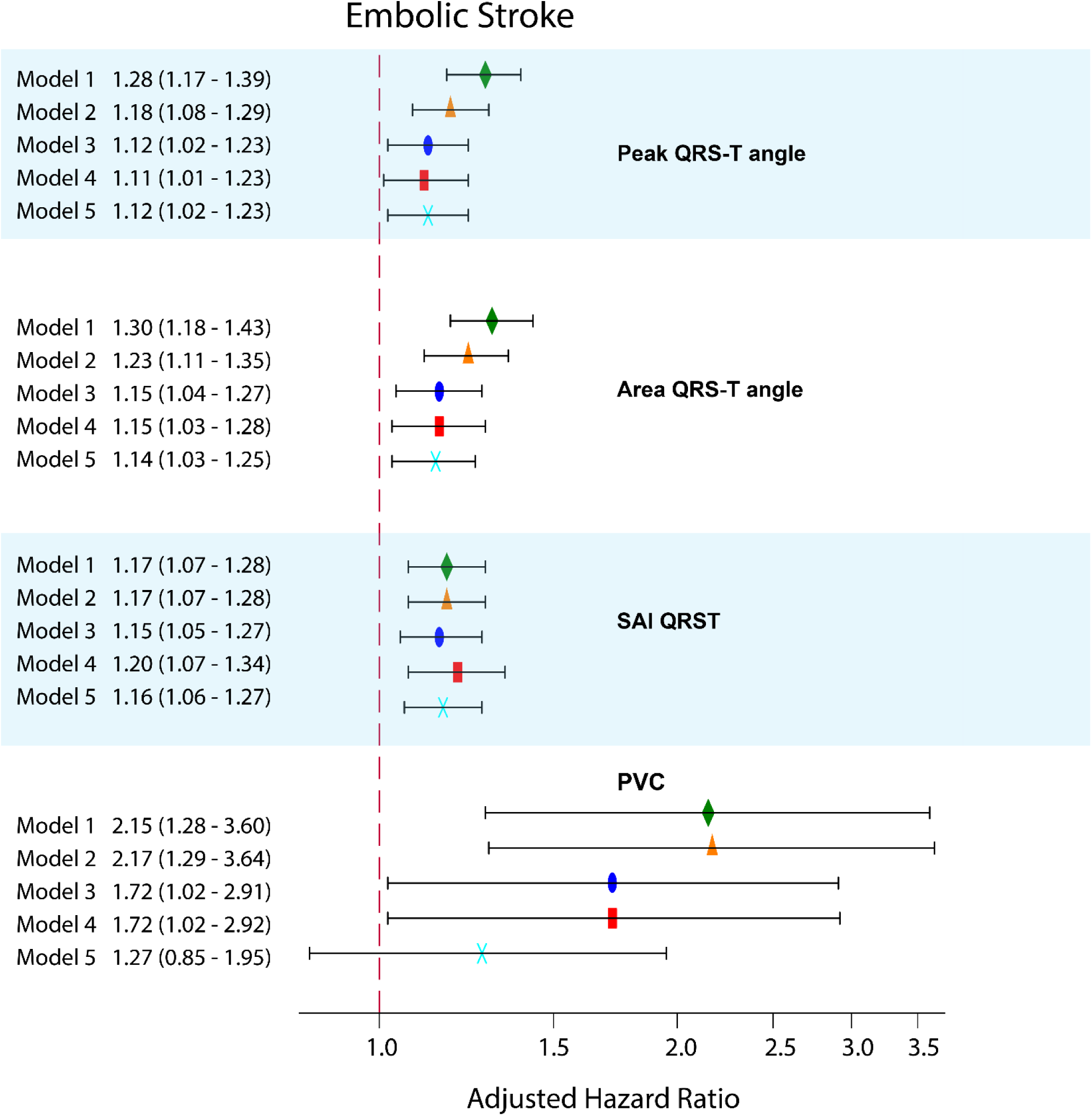
Adjusted Cox proportional hazard ratio (HR) with 95% confidence interval (CI) for the association of peak and area QRS-T angle, SAI QRST, and a PVC on a 10-second ECG with EBI. Model 1 (green diamond) was adjusted for age, sex, and race-study center group. Model 2 (orange triangle) in addition to model 1 covariates included BMI, WHR, total cholesterol, HDL, triglyceride level, use of lipid-lowering medications, current smoking, consumption of alcohol, level of physical activity at leisure time, diabetes, hypertension, levels of systolic and diastolic blood pressure, use of blood pressure-lowering drugs, eGFRcKD-EPI, and the presence of carotid artery plaque. Model 3 (blue oval) in addition to model 2 covariates included abnormal P wave indices (P axis, PR interval, PTFv1), heart rate, use of antiarrhythmic drugs, anticoagulants, and aspirin, the presence of PACs, S or V median beat at any visit and AF at baseline or at any time during follow-up. Model 4 (red rectangle) adjusted for model 3 covariates plus QRS duration, QTc interval, presence of BBB/IVCD, ECG-LVH, categorical GEH risk score, and the presence of PVCs. Time-updated model 5 (cyan X) included all model 4 covariates with the following ECG variables that were updated at the date of ECG recording in visits 1-5: QRS duration, QTc interval, presence of BBB/IVCD, ECG-LVH, GEH variables, abnormal P-axis, PTFv1 and PR interval, heart rate, presence of PAC and PVC on a single 10-second 12-lead ECG, and a type of median beat. Black lines correspond to 95% CI bounds.

### The outcomes: incident stroke subtypes

The follow-up procedures of ARIC study participants^28^ have been described previously. In this study, we included the follow-up period from January 1st, 1987, to December 31st, 2018, for participants enrolled in Forsyth County, Minneapolis, and Washington County field centers. The follow-up period from January 1st, 1987, to December 31st, 2017, was included for participants enrolled in the city of Jackson field center. Methods for the ascertainment of stroke events have been previously described.^29^ Briefly, records for possible stroke-related hospitalizations were selected based on the International Classification of Diseases, 9^th^ Revision *(ICD-9)* codes 430-438 until 1997, *ICD-9* codes 430-436 and *ICD-10* codes G45.X, I60.X, I61.X, I62.X, I63.X, I65.X, I66.X, and I67.X afterward. Data on fatal stroke was collected through linkage with the National Death Index. Strokes were then identified by a computer algorithm and adjudicated by physician reviewers.^29^

In this study, we included three incident stroke subtype outcomes: a first definite or probable (1) thrombotic brain infarction (TBI), (2) non-carotid cardioembolic brain infarction (EBI), and (3) subarachnoid or intracranial brain hemorrhage (intracerebral hemorrhage, ICH). If a stroke case met the criteria for two different diagnostic categories, the following hierarchy was used: hemorrhagic above cardioembolic, above thrombotic, as previously described.^29^ In this study, we considered multiple stroke events for the same participant. Thus, a participant could develop all three incident stroke subtype outcomes as three separate events.

### Atrial fibrillation, ectopic atrial complexes, and use of anticoagulants/aspirin

Prevalent AF included AF or atrial flutter diagnosed on the 1^st^ ECG. Incident AF included AF detected on follow-up 12-lead ECG or hospital discharge records and death certificates with *ICD-9-CM* code 427.31 or 427.32 or *ICD-10* code I48 listed in any position.^30, 31^

Premature atrial complexes (PACs) were identified on 10-second 12-lead ECG, as described above.

The ARIC study participants were asked to bring their medications to the clinic, where ARIC staff filled out the Medication Survey Form (based partly on self-report by the participants). The use of anticoagulant and aspirin-containing medications in the past two weeks was self-reported and validated by medication inventory at every study visit.

### Measures of an atrial substrate and use of antiarrhythmic medications

The atrial substrate was characterized by the following P-wave indices: frontal P axis, P-terminal force in lead V_1_ (PTF_V1_), and PR interval duration. All P-wave indices, including amplitude and duration of the terminal negative phase (P-prime) of biphasic (positive/negative) P wave in lead V1, were automatically measured by the 12SL algorithm (GE Marquette Electronics, Milwaukee, WI). PTF_V1_ was calculated as a product of P-prime amplitude and duration.^32, 33^

The use of antiarrhythmic medications included in the study was self-reported and subsequently validated by medication inventory. Antiarrhythmic medications included class I, II (beta-blockers), III, IV (phenylalkylamines, and benzothiazepines calcium channel blockers), or V (digoxin) antiarrhythmic agents.

### Carotid artery plaque

Carotid artery intima-media thickness was measured in three locations of the carotid arteries bilaterally; at the extracranial carotid artery 1 cm proximal to the dilatation of the carotid bulb, the carotid bifurcation 1 cm proximal to the flow divider, and the internal carotid artery 1 cm distal to the flow divider, using high-resolution B-mode ultrasound (Biosound 2000 II SA; Biosound, Indianapolis, IN), as previously described.^34-36^ ARIC ultrasound readers considered an atherosclerotic plaque to be present at any of the six segments if wall thickness exceeded 1.5 mm, or if there was a lumen encroachment or irregular intimal surface or image characteristics indicative of structural heterogeneity of the arterial wall.

### Prevalent cardiovascular disease and common cardiovascular risk factors

The baseline prevalence of cardiovascular disease (CVD) was defined as the presence of prevalent coronary heart disease (CHD), peripheral artery disease (PAD), or heart failure (HF) at the first visit. Prevalent CHD included a self-reported physician-diagnosed heart attack, baseline ECG evidence of myocardial infarction (MI) by the Minnesota code^37^, or a history of coronary revascularization (either via coronary artery bypass surgery or percutaneous coronary intervention). Prevalent PAD was defined as self-reported history of leg pain during walking that disappeared within 10 minutes after rest, an ankle-brachial index <0.9, or leg artery revascularization.^38^ Prevalent HF was defined as asymptomatic (stage 3 by the Gothenburg criteria) HF, manifesting by cardiac and pulmonary symptoms, on medical treatment^39^, or self-reported use of HF medication.

Body mass index (BMI) was calculated as weight (kg)/height (m)^2^. Waist-to-hip ratio (WHR) was calculated as a measure of fat distribution. During the clinic visit, sitting blood pressure was measured three times, each after 5 minutes of rest. The average of the second and third of 3 consecutive measurements was used to calculate systolic and diastolic blood pressure levels. Hypertension was defined as blood pressure of ≥140/90 mm Hg, or self-reported use of antihypertensive drugs. Self-reported use of antihypertensive and lipid-lowering medications in the past two weeks was validated by medication inventory at every study visit. Diabetes was defined as nonfasting blood glucose ≥200 mg/dL, fasting blood glucose ≥126 mg/dL, self-reported physician diagnosis of diabetes, or self-reported use of drugs to treat diabetes. Kidney function was assessed by estimated glomerular filtration rate (eGFR) calculated using the CKD Epidemiology Collaboration equation (CKD-EPI).^40^ Physical activity was measured during leisure time, using the semicontinuous indices ranging from 1 (low) to 5 (high), defined by modified Baecke questionnaire.^41^ Participants self-reported smoking and consumption of alcoholic beverages.

### Statistical analyses

Continuous variables were presented as means and standard deviation (SD). We used ANOVA (for normally distributed continuous variables) and *χ*^2^ test (for categorical variables) to compare baseline clinical characteristics in participants with 0 - 5 abnormal GEH parameters.

#### Analysis of circular variables

An unadjusted comparison of circular variables (spatial QRS-T angle, SVG azimuth, and SVG elevation) in participants with different types of stroke was performed using the Wheeler-Watson-Mardia test.

Because distributions of QRS-T and SVG elevation angles were normal or nearly normal, we included them in all conventional statistical analyses without transformation. The SVG azimuth angle is expressed as an axial variable, ranging from −180° to +180°. Thus, as recommended^42^, we transformed SVG azimuth by doubling its value and then adding 360°. We then analyzed the SVG azimuth in multivariable Cox regression analysis and converted it back for interpretation.

#### Survival analyses

We built Cox proportional hazard risk models. All continuous ECG exposure variables were expressed as their z score to standardize comparisons. The proportional-hazards assumption was verified using *stcox* PH-assumptions suite of tests implemented in STATA (StataCorp LP, College Station, TX), and exceptions were reported. To adjust for confounders, we constructed five models with incremental adjustment. Model 1 was adjusted for demographic characteristics (age, sex, and race-study center group).

Model 2, in addition to model 1 covariates, was adjusted for prevalent at baseline CVD and risk factors (BMI, WHR, total cholesterol, HDL, triglyceride level, use of lipid-lowering medications, current smoking, consumption of alcohol, level of physical activity at leisure time, diabetes, hypertension, levels of systolic and diastolic blood pressure, use of blood pressure-lowering drugs, eGFR_CKD-EPI_, and the presence of carotid artery plaque).

Model 3 further added characteristics of an atrial substrate [abnormal P axis (less than 0 or above 75°)^6^, abnormal PR interval (less than 120 ms or above 200 ms)^31^, abnormal PTF_V1_ (≤ −4000 μV*ms)^6^], heart rate, use of antiarrhythmic drugs, anticoagulants, and aspirin. In cases of P-wave absence, PR interval and P-wave indices were categorized as abnormal. Model 3 also added presence of PACs on 10-second 12-lead ECG at any visit, S or V median beat at any visit, and presence of AF at baseline or at any time during follow-up (assuming incident AF has been present since baseline).^43^

Model 4 adjusted for model 3 covariates plus characteristics of a ventricular substrate (QRS duration, QTc interval, presence of BBB/IVCD, ECG-LVH, categorical GEH risk score, and presence of PVCs on 10-second 12-lead ECG at any visit). Categorical GEH risk score was not included in the models with continuous GEH variables serving as predictors.

Time-updated model 5 included all model 4 covariates, as well as ECG variables that were updated at the date of ECG recording in visits 1-5, as time-updated exposure (QRS duration, QTc interval, presence of BBB/IVCD, ECG-LVH, and GEH variables), and time-updated covariates (abnormal P-axis, abnormal PTF_V1_, abnormal PR interval, heart rate, presence of PAC and PVC on a single 10-second 12-lead ECG, and a type of median beat).

Associations of continuous ECG variables with stroke were also studied using adjusted (model 1) Cox regression models incorporating cubic splines with 4 knots.

#### Sensitivity analyses

To assess the robustness of the study findings, we conducted several sensitivity analyses. First, we repeated all survival analyses after excluding participants with two or three different types of stroke. Also, as statistical power for hemorrhagic stroke was less than for ischemic stroke, we constructed sequential (instead of incremental) sets of models 3-5 with hemorrhagic stroke outcome, to avoid overfitting. Model 3 included age, sex, race and study center, prevalent at baseline CVD, abnormal P axis, abnormal PR interval, abnormal PTF_V1_, heart rate, use of antiarrhythmic drugs, anticoagulants, and aspirin, presence of PACs on 10-second 12-lead ECG at any visit, S or V median beat at any visit and presence of AF at baseline or at any time during follow-up. Model 4 adjusted for age, sex, race and study center, QRS duration, QTc interval, presence of BBB/IVCD, ECG-LVH, presence of PVCs on 10-second 12-lead ECG at any visit, and categorical GEH risk score. Categorical GEH risk score was not included in the models with continuous GEH variables serving as predictors. Time-updated model 5 included age, sex, race and study center, and ECG variables that were updated at the date of ECG recording in visits 1-5 (presence of TD-IBBB on a single 10-second 12-lead ECG, GEH variables, QRS duration, QTc interval, presence of BBB/IVCD on a single 10-second 12-lead ECG, ECG-LVH, abnormal P-axis, abnormal PTFV1, abnormal PR interval, heart rate, presence of PAC and PVC on a single 10-second 12-lead ECG, and a type of median beat).

Statistical analyses were performed using STATA MP 16.1 (StataCorp LP, College Station, TX). A *P* value < 0.05 was considered statistically significant.

## Results

### Study population

More than half of the participants had 0-1 abnormal GEH measures, and only 6% had 4-5 abnormal GEH metrics (Table 1). PVCs were detected on a 12-lead ECG in 280 (1.93%) participants at baseline and 1,202 (8.30%) participants at any visit. TD-IBBB was observed in at least one ECG in 122(0.84%) participants. Participants with more abnormal GEH were more likely to have prevalent CVD and traditional cardiovascular risk factors (hypertension, diabetes), and use antiarrhythmic medications and anticoagulants. Of note, there was a gradual increase in the presence of carotid plaque and a gradual decrease in the level of HDL, consistent with a growth in the number of abnormal GEH metrics (Table 1). As expected, abnormal P wave indices, PACs, PVCs, and ventricular conduction abnormalities were more likely to be observed in participants with abnormal GEH.

**Table 1.** Comparison of baseline clinical and ECG characteristics in participants with abnormal GEH measurements

| Characteristics | Normal GEH<br>(n=2,837; 19.5%) | 1 abnormal GEH<br>(n=5,290; 36.6%) | 2 abnormal GEH<br>(n=4,090; 28%) | 3 abnormal GEH<br>(n=1,382; 10%) | 4 abnormal GEH<br>(n=555; 4%) | 5 abnormal GEH<br>(n=325; 2%) | ANOVA<br>or $\chi^2$ *P* |
| --- | --- | --- | --- | --- | --- | --- | --- |
| Age(SD), y | 53.4(5.7) | 54.0(5.7) | 54.3(5.8) | 54.8(5.8) | 55.1(5.8) | 56.8(5.6) | <0.0001 |
| Female, n(%) | 2,777(97.9) | 2,872(54.3) | 1,598(39.1) | 527(38.1) | 113(20.4) | 117(36.0) | <0.0001 |
| White, n(%) | 2,189(77.2) | 4,075(77.0) | 3,159(77.2) | 979(70.8) | 340(61.3) | 193(59.4) | <0.0001 |
| Prevalent cardiovascular disease, n(%) | 291(10.3) | 572(10.8) | 503(12.3) | 235(17.0) | 126(22.7) | 107(32.9) | <0.0001 |
| Body mass index(SD), kg/m<sup>2</sup> | 27.7(5.6) | 27.8(5.3) | 27.4(5.0) | 27.3(5.2) | 26.9(4.6) | 27.7(4.9) | 0.0001 |
| Waist-to-hip ratio (SD) | 0.896(0.081) | 0.929(0.077) | 0.934(0.075) | 0.934(0.077) | 0.943(0.069) | 0.940(0.070) | <0.0001 |
| Hypertension, n(%) | 739(26.1) | 1,709(32.3) | 1,391(34.0) | 569(41.2) | 281(50.6) | 176(54.2) | <0.0001 |
| Antihypertensive drugs, n(%) | 750(26.4) | 1,525(28.8) | 1,153(28.2) | 484(35.0) | 230(41.4) | 145(44.6) | <0.0001 |
| Systolic blood pressure(SD), mmHg | 116.4(16.5) | 120.2(17.6) | 121.8(18.3) | 124.9(20.0) | 129.8(24.7) | 129.8(23.4) | <0.0001 |
| Diastolic blood pressure(SD), mmHg | 70.8(9.9) | 73.4(10.6) | 74.0(10.8) | 74.9(12.4) | 78.0(14.6) | 77.8(14.7) | <0.0001 |
| eGFR<sub>CKD-EPI</sub> (SD), mL/min/1.73 m<sup>2</sup> | 96.9(14.3) | 95.7(15.1) | 95.3(14.9) | 94.4(16.3) | 94.4(17.3) | 90.2(18.8) | <0.0001 |
| Diabetes, n(%) | 247(8.7) | 556(10.5) | 477(11.7) | 195(14.1) | 98(17.7) | 64(19.7) | <0.0001 |
| Current tobacco smoker, n(%) | 647(22.8) | 1,315(24.9) | 1,071(26.2) | 426(30.8) | 186(33.5) | 85(26.2) | <0.0001 |
| Current alcohol drinker, n(%) | 1,519(53.5) | 3,044(57.5) | 2,390(58.4) | 780(56.4) | 313(56.4) | 170(52.3) | 0.001 |
| Leisure physical activity score(SD) | 2.42(0.58) | 2.37(0.57) | 2.37(0.56) | 2.33(0.58) | 2.27(0.57) | 2.27(0.56) | <0.0001 |
| Total cholesterol(SD), mmol/L | 5.66(1.09) | 5.54(1.08) | 5.50(1.06) | 5.51(1.05) | 5.55(1.11) | 5.53(1.07) | <0.0001 |
| HDL cholesterol(SD), mg/dL | 57.2(16.7) | 51.3(17.2) | 49.8(16.7) | 49.7(17.0) | 48.3(17.1) | 23.2(25.7) | <0.0001 |
| Triglycerides(SD), mmol/L | 1.39(0.97) | 1.50(1.00) | 1.50(1.03) | 1.53(1.07) | 1.53(1.00) | 1.55(1.01) | <0.0001 |
| Lipid-lowering drugs, n(%) | 78(2.8) | 148(2.8) | 123(3.0) | 46(3.3) | 17(3.1) | 15(4.6) | 0.448 |
| Plaque in any carotid site, n(%) | 688(24.3) | 1,701(32.2) | 1,435(35.1) | 533(38.6) | 243(43.8) | 152(46.8) | <0.0001 |
| Aspirin, n(%) | 1,389(49.0) | 2,466(46.6) | 1,905(46.6) | 637(46.1) | 234(42.2) | 147(45.2) | 0.054 |
| Anticoagulants, n(%) | 2(0.07) | 12(0.23) | 16(0.39) | 11(0.80) | 7(1.26) | 9(2.77) | <0.0001 |
| Antiarrhythmic drugs, n(%) | 293(10.3) | 668(12.6) | 566(13.8) | 253(18.3) | 132(23.8) | 79(24.3) | <0.0001 |
| Heart rate(SD), bpm | 67.7(9.6) | 66.1(9.9) | 65.2(10.3) | 66.3(11.4) | 65.5(10.6) | 67.3(11.0) | <0.0001 |
| QRS duration(SD), ms | 85.7(7.8) | 90.4(9.5) | 93.8(10.5) | 98.7(14.9) | 107.7(20.2) | 106.5(25.5) | <0.0001 |
| Bazett-corrected QTc(SD), ms | 417.8(18.3) | 414.7(18.1) | 414.3(17.8) | 418.2(20.2) | 423.1(24.3) | 428.3(31.7) | <0.0001 |
| ECG-LVH, n(%) | 16(0.6) | 109(2.1) | 170(4.2) | 197(14.3) | 195(35.1) | 119(36.6) | <0.0001 |
| BBB/IVCD, n(%) | 30(1.1) | 111(2.1) | 146(3.6) | 138(10.0) | 123(22.2) | 60(18.5) | <0.0001 |
| Any visit TD-IBBB, n(%) | 16(0.56) | 54(1.02) | 37(0.90) | 9(0.65) | 2(0.36) | 4(1.23) | 0.181 |
| Any visit PVC, n(%) | 152(5.4) | 402(7.6) | 388(9.5) | 166(12.0) | 61(11.0) | 33(10.2) | <0.0001 |
| Any visit PAC, n(%) | 202(7.1) | 460(8.7) | 361(8.8) | 129(9.3) | 44(7.9) | 29(8.9) | 0.097 |
| Any visit Atrial Fibrillation, n(%) | 472(16.6) | 1,136(21.5) | 897(21.9) | 357(25.8) | 128(23.1) | 47(14.5) | <0.0001 |
| Abnormal PR interval, n(%) | 219(7.7) | 447(8.5) | 367(9.0) | 142(10.3) | 80(14.4) | 47(14.5) | <0.0001 |
| Abnormal Frontal P axis, n(%) | 188(6.6) | 399(7.5) | 345(8.4) | 152(11.0) | 67(12.1) | 37(11.4) | <0.0001 |
| Abnormal PTF in V1, n(%) | 171(6.0) | 439(8.3) | 440(10.8) | 217(15.7) | 119(21.4) | 60(18.5) | <0.0001 |
| Median beat S or V at any visit, n(%) | 44(1.6) | 140(2.7) | 134(3.3) | 63(4.6) | 24(4.3) | 16(4.9) | <0.0001 |

HDL=High-density lipoprotein; SD=standard deviation

### Incident stroke

Over a median follow-up of 24.5 years, there were 899 thrombotic strokes (incidence 2.87; 95% CI 2.69-3.07 per 1000 person-years), 400 cardioembolic strokes (incidence 1.26; 95% CI 1.14-1.39 per 1000 person-years), and 187 hemorrhagic strokes (incidence 0.59; 95% CI 0.51-0.68 per 1000 person-years). Two participants experienced all three types of incident stroke; 62 participants suffered from both types of ischemic stroke (TBI and EBI), and another 17 participants endured both ICH and either TBI or EBI. Overall, baseline GEH was more abnormal in participants who developed any type of stroke during follow-up (Table 2).

**Table 2.** GEH in participants with different types of stroke

| Characteristics | No stroke<br>(n=13,076) | Only TS (n=819) | Only ES (n=335) | Only HS (n=168) | TS & ES<br>(n=62) | HS & TS/ES<br>(n=19) | P-value |
| --- | --- | --- | --- | --- | --- | --- | --- |
| Heart rate(SD), bpm | 66.1(10.1) | 66.9(10.9) | 66.4(10.9) | 66.0(10.9) | 67.8(10.6) | 67.6(5.9) | 0.282 |
| QRS duration(SD), ms | 92.2(12.4) | 93.8(13.8) | 93.0(11.1) | 90.7(11.1) | 91.1(11.5) | 91.4(11.8) | 0.004 |
| Bazett QTc(SD), ms | 415.9(19.0) | 418.8(20.5) | 418.5(19.1) | 417.4(20.3) | 418.4(16.6) | 416.4(19.2) | 0.0002 |
| ECG-LVH, n(%) | 674(5.2) | 79(9.7) | 34(10.2) | 13(7.7) | 3(4.8) | 3(15.8) | <0.0001 |
| PVC any visit, n(%) | 1,049(8.0) | 87(10.6) | 53(15.8) | 8(4.8) | 4(6.5) | 1(5.3) | <0.0001 |
| BBB/IVCD, n(%) | 541(4.1) | 42(5.1) | 15(4.5) | 7(4.2) | 2(3.2) | 1(5.3) | 0.829 |
| TD-IBBB any visit, n(%) | 109(0.83) | 7(0.85) | 2(0.60) | 3(1.79) | 1(1.61) | 0(0) | 0.754 |
| Atrial fibrillation any visit, n(%) | 2,549(19.5) | 183(22.3) | 233(70.0) | 29(17.3) | 38(61.3) | 5(26.3) | <0.0001 |
| PAC any visit, n(%) | 1,079(8.3) | 82(10.0) | 43(12.8) | 15(8.9) | 5(8.1) | 1(5.3) | 0.037 |
| Median beat S or V at any visit, n(%) | 352(2.7) | 14(1.7) | 47(14.0) | 2(1.2) | 5(8.1) | 1(5.3) | <0.0001 |
| Abnormal PR interval, n(%) | 1,159(8.9) | 82(10.0) | 40(11.9) | 11(6.6) | 9(14.5) | 1(5.3) | 0.121 |
| Abnormal Frontal P axis, n(%) | 1,065(8.1) | 76(9.3) | 27(8.1) | 11(6.6) | 6(9.7) | 3(15.8) | 0.613 |
| Abnormal PTF in V1, n(%) | 1,240(9.5) | 125(15.3) | 47(14.0) | 19(11.3) | 9(14.5) | 6(31.6) | <0.0001 |
| Peak QRS-T angle(circular 95%CI), ° | 41.5(41.0-42.0) | 46.4(44.3-48.6) | 49.0(45.3-52.7) | 48.0(43.1-52.9) | 47.7(38.8-56.6) | 56.9(35.4-78.5) | <0.0001 |
| Area QRS-T angle(circular 95%CI), ° | 60.0(59.6-60.5) | 63.8(61.7-65.9) | 65.3(62.0-68.6) | 61.9(57.7-66.2) | 65.0(57.9-72.1) | 69.3(52.7-85.9) | 0.442 |
| Peak SVG elevation(circular SD), ° | 63.1(62.9-63.4) | 64.9(63.9-66.0) | 66.2(64.5-67.9) | 63.3(61.0-65.5) | 66.5(62.8-70.2) | 66.7(58.0-75.4) | 0.006 |
| Area SVG elevation(circular SD), ° | 67.2(66.9-67.5) | 69.2(68.0-70.4) | 70.0(68.0-72.0) | 67.7(65.3-70.1) | 69.3(64.7-74.0) | 70.5(61.2-80.0) | 0.041 |
| Peak SVG azimuth(circular SD), ° | 2.4(2.1-2.8) | 2.4(0.9-3.9) | 4.1(1.5-6.7) | 6.8(3.5-10.1) | 4.7(-0.5 to 9.9) | 4.8(-6.6 to 16.2) | 0.095 |
| Area SVG azimuth(circular SD), ° | 24.5(24.1-24.9) | 24.6(22.9-26.3) | 23.4(20.8-26.0) | 27.6(24.1-31.0) | 27.2(21.6-32.8) | 25.1(14.0-36.3) | 0.842 |
| SAI QRST, mV*ms | 142.2(49.9) | 149.5(55.9) | 151.1(55.5) | 146.4(50.5) | 143.0(47.9) | 157.8(58.1) | <0.0001 |
| Peak SVG magnitude(SD), mV | 1.60(0.44) | 1.66(0.46) | 1.64(0.49) | 1.68(0.48) | 1.66(0.41) | 1.62(0.39) | 0.0002 |
| SVG magnitude, mV | 1.70(0.48) | 1.76(0.53) | 1.74(0.52) | 1.81(0.56) | 1.72(0.50) | 1.79(0.43) | 0.003 |

### Ventricular substrate and cardioembolic stroke

In minimally adjusted model 1, nearly all measures of the ventricular substrate (except QRS duration, SVG magnitude, and ventricular conduction abnormalities) associated with incident cardioembolic stroke (Table 3). Model 2 showed that CVD and cardiovascular risk factors explained a significant portion of an association of QRS-T angle, SVG direction, QTc interval, and ECG-LVH with incident EBI (Figure 2). Of note, an association of SAI QRST and PVCs with cardioembolic stroke was independent of CVD and cardiovascular risk factors. Neither persistent BBB/IVCD nor TD-IBBB associated with cardioembolic stroke.

**Table 3.**
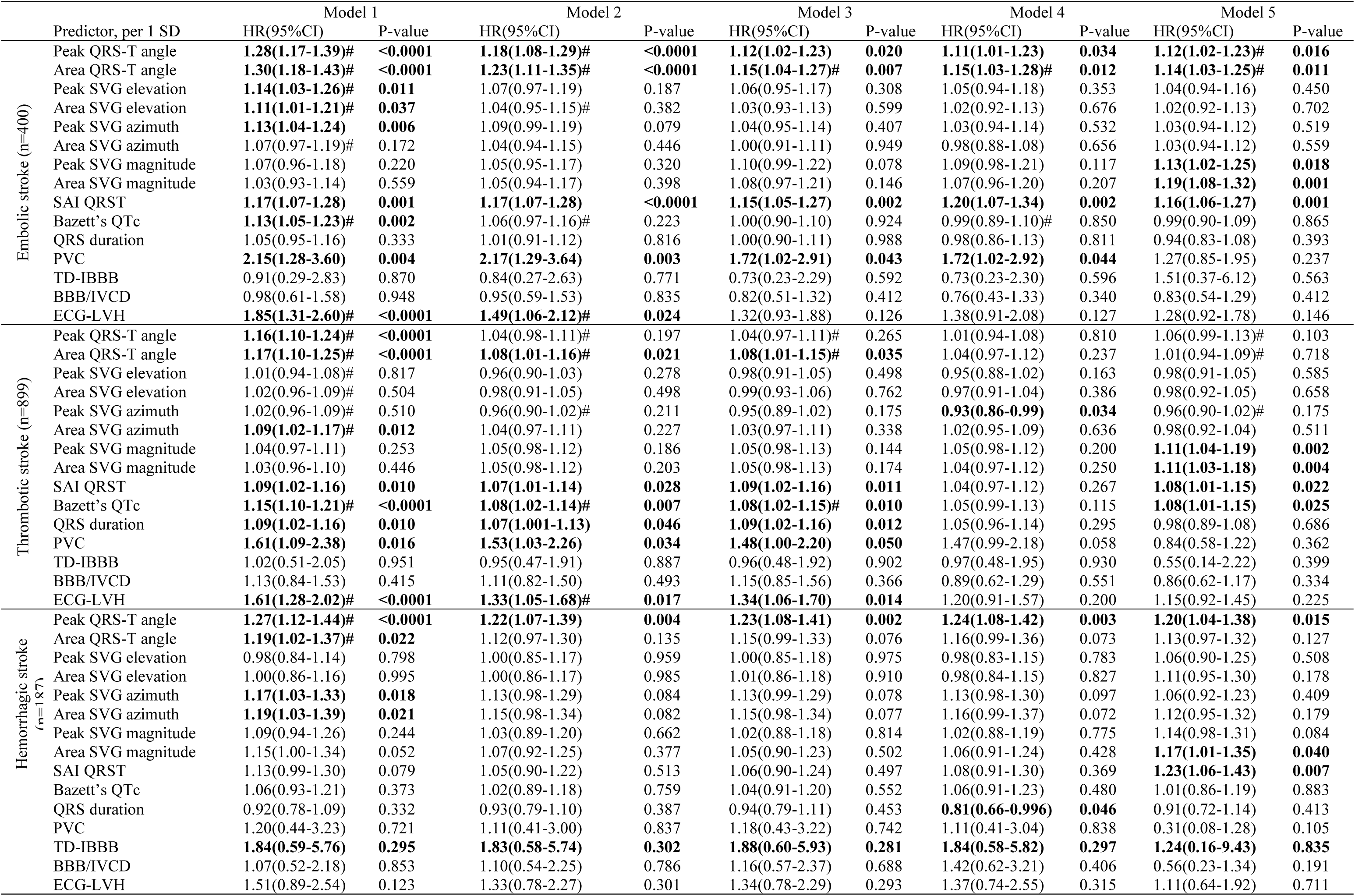
Association of GEH with incident stroke in Cox models.

The addition of the atrial substrate metrics in model 3 resulted in further attenuation of the association of QRS-T angle and PVCs with EBI. It thoroughly explained the association of SVG direction and traditional ECG metrics (QTc, ECG-LVH) with cardioembolic stroke. Model 4 confirmed that spatial QRS-T angle, SAI QRST, and PVCs represent specific features of the ventricular substrate independently associated with EBI. Notably, the association of QRS-T angle and SAI QRST with EBI was “dose-dependent,” as demonstrated by the gradually increasing risk of EBI across the distribution of QRS-T angle and SAI QRST (Figure 3). In time-updated model 5, SVG magnitude emerged as significant independent transient exposure associated with incident cardioembolic stroke (Table 3). SAI QRST and QRS-T angle remained significantly associated, too (Figure 2).

**Figure 3.**
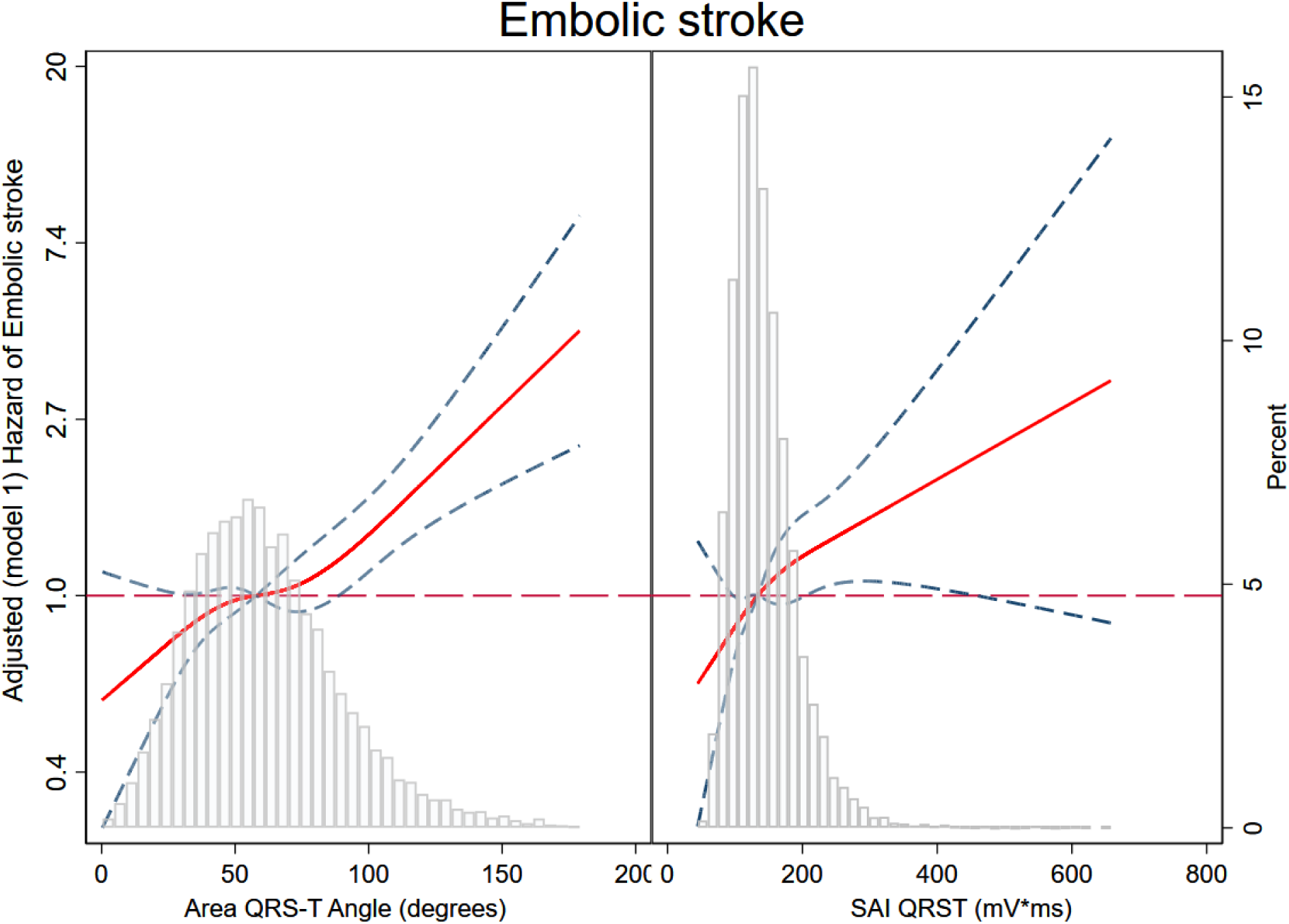
Adjusted (model 1) risk of cardioembolic brain infarction associated with area QRS-T angle (A) and SAI QRST (B). Restricted cubic spline with 95% CI shows a change in the hazard ratio (Y-axis) in response to QRS-T angle (A) and SAI QRST (B) change (X-axis). 50thpercentile of QRS-T angle (A) and SAI QRST (B) is selected as a reference. Knots of area QRS-T angle are at 21 – 48 – 69 – 114 degrees. Knots of SAI QRST are at 83 – 119 – 151 – 230 mV*ms.

Sensitivity analysis in participants with pure EBI outcome (Table 4) strengthened observed findings, reporting higher point estimates, especially for time-dependent SVG magnitude in model 5.

**Table 4.**
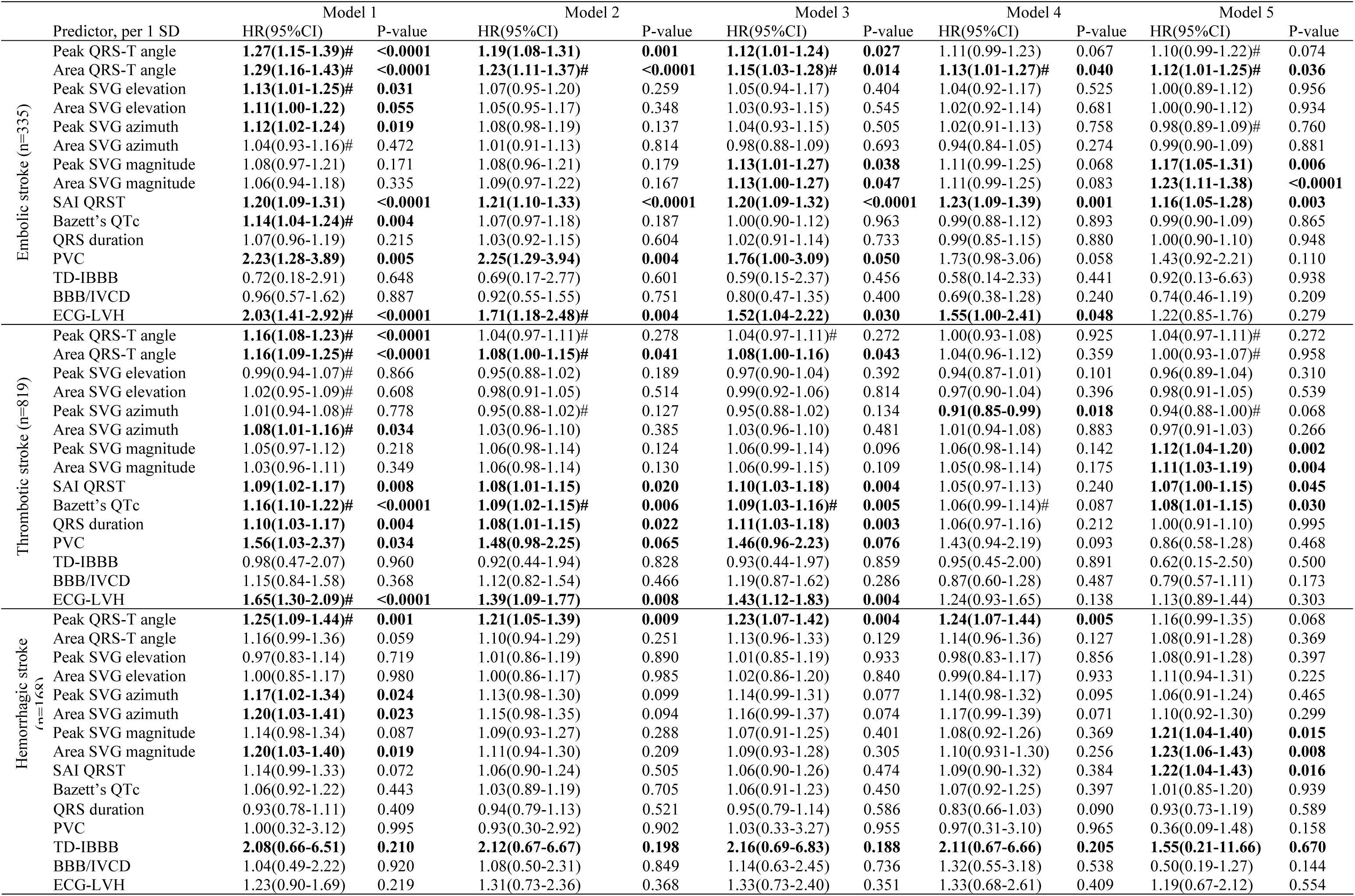
Association of GEH with incident stroke in Cox models with pure stroke type outcomes.

### Ventricular substrate and thrombotic stroke

In minimally adjusted model 1, nearly all traditional and novel measures of the ventricular substrate (except SVG elevation, magnitude, and ventricular conduction abnormalities) associated with TBI, although the strength of association was weaker for thrombotic than cardioembolic stroke (Table 3). Similar to EBI, the risk of TBI increased across the distribution of the QRS-T angle and SAI QRST (Figure 4). Adjustment for CVD and cardiovascular risk factors in model 2 substantially attenuated the association for all ECG metrics. Spatial QRS-T angle, SAI QRST, QTc, and QRS duration, PVCs, and ECG-LVH remained associated with TBI in model 2 (Figure 5).

**Figure 4.**
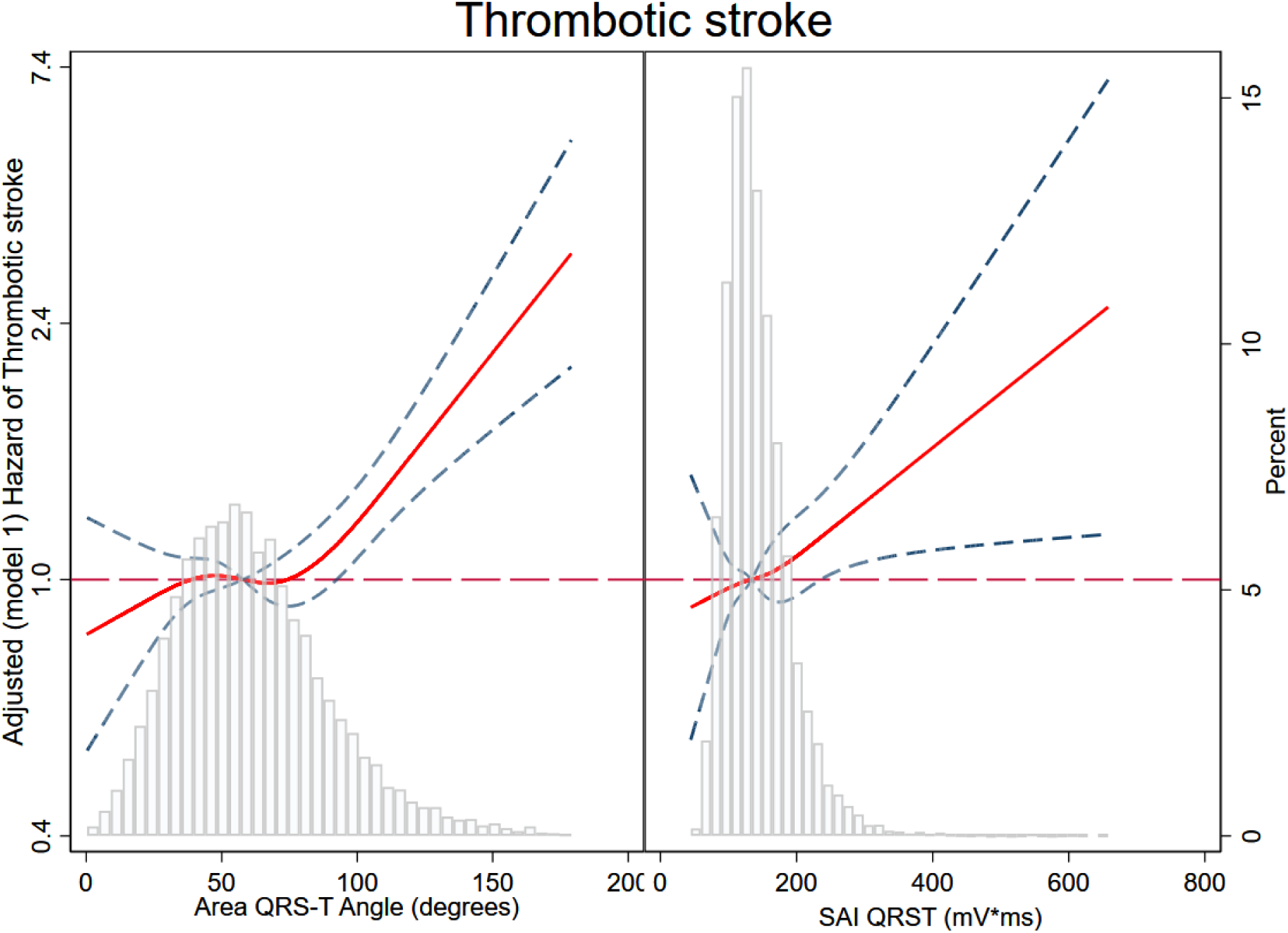
Adjusted (model 1) risk of thrombotic brain infarction associated with area QRS-T angle (A) and SAI QRST (B). See Figure 3 legend for the details.

**Figure 5.**
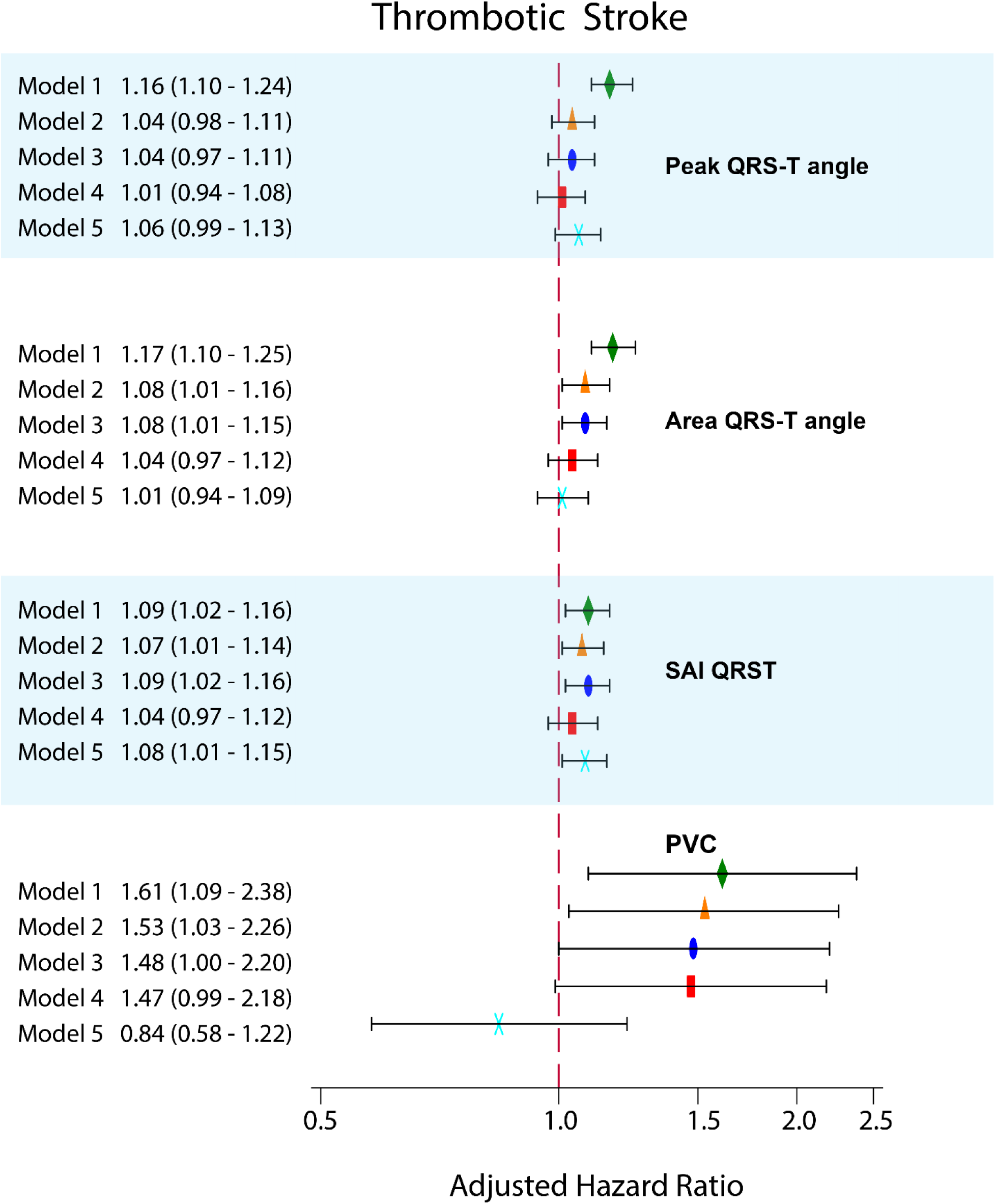
Adjusted Cox proportional hazard ratio (HR) with 95% confidence interval (CI) for the association of peak and area QRS-T angle, SAI QRST, and a PVC on a 10-second ECG with TBI. See Figure 2 legend for the details.

However, in contrast to cardioembolic stroke, further adjustment for atrial substrate in model 3 did not change the strength of associations (Table 3), suggesting that an association of a ventricular substrate with TBI was independent of atrial substrate. Again, in contrast to the cardioembolic stroke, model 4 showed that there was no independent association of GEH or other measures of the ventricular substrate with incident thrombotic stroke, as previously observed in models 1-3 associations attenuated and were not statistically significant.

Interestingly, time-updated model 5 revealed a strengthened association of SAI QRST, QT interval, and SVG magnitude with TBI, suggesting the importance of time-dependent transient ventricular substrate (Table 3).

Sensitivity analysis with pure TBI outcome demonstrated identical results, supporting the robustness of the study findings.

### Ventricular substrate and hemorrhagic stroke

As ICH outcome was less frequent than ischemic stroke outcomes, resulting in a lesser statistical power, sensitivity analyses with the ICH outcome highlighted significant observations (Tables 4-5). In contrast to ischemic stroke, very few features of ventricular substrate associated with incident hemorrhagic stroke. In model 1, SVG azimuth and magnitude were associated with ICH. Still, their association was explained mainly by CVD and cardiovascular risk factors, as shown by attenuated hazard ratio estimates for SVG azimuth and magnitude in model 2 (Tables 3-4). Similar to ischemic stroke, the risk of ICH was increasing across the distribution of the spatial area QRS-T angle and SAI QRST (Figure 6), but the strength of association was weaker and did not reach statistical significance.

**Table 5.**
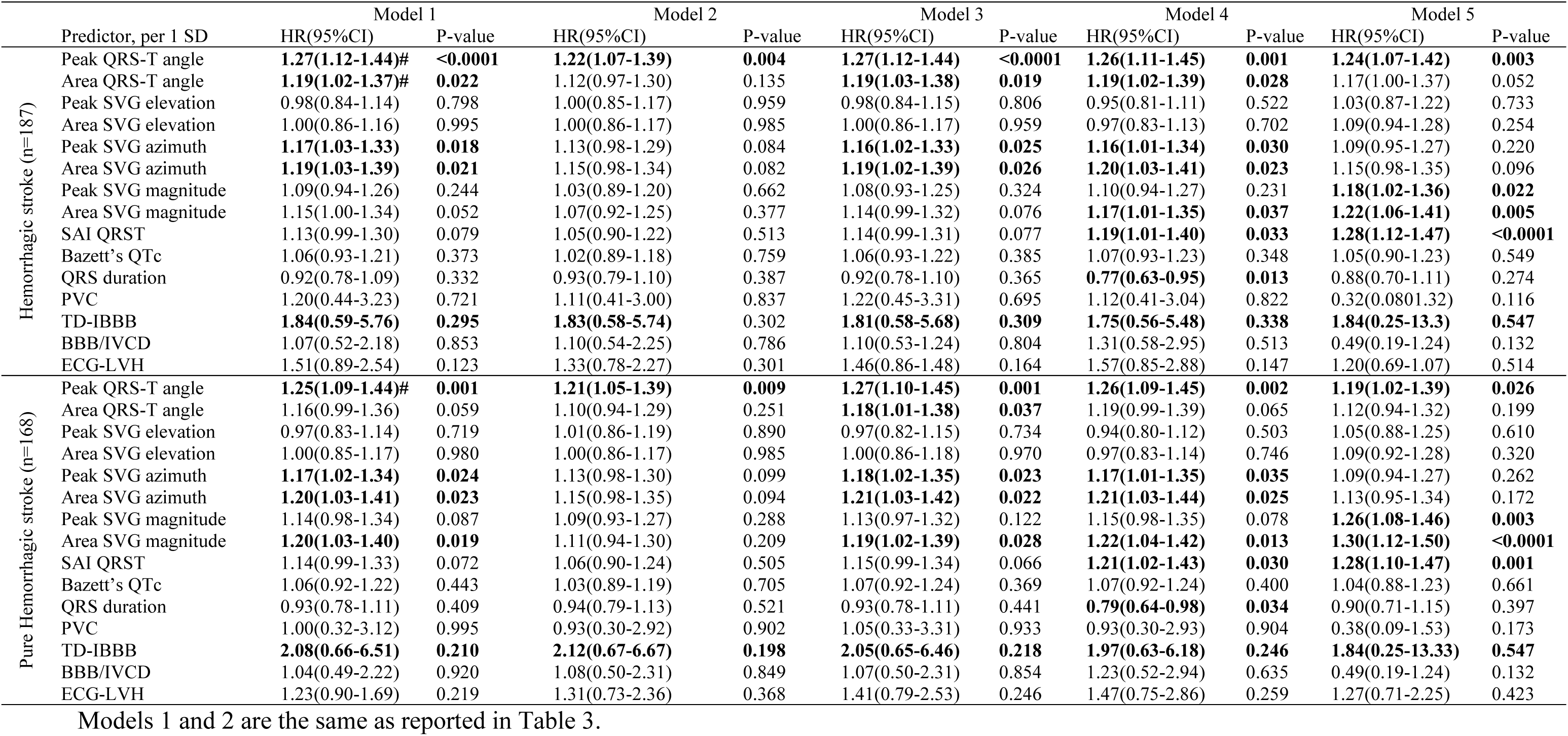
Association of GEH with incident hemorrhagic stroke in Cox models with sequential adjustment applied to models 3-5.

**Figure 6.**
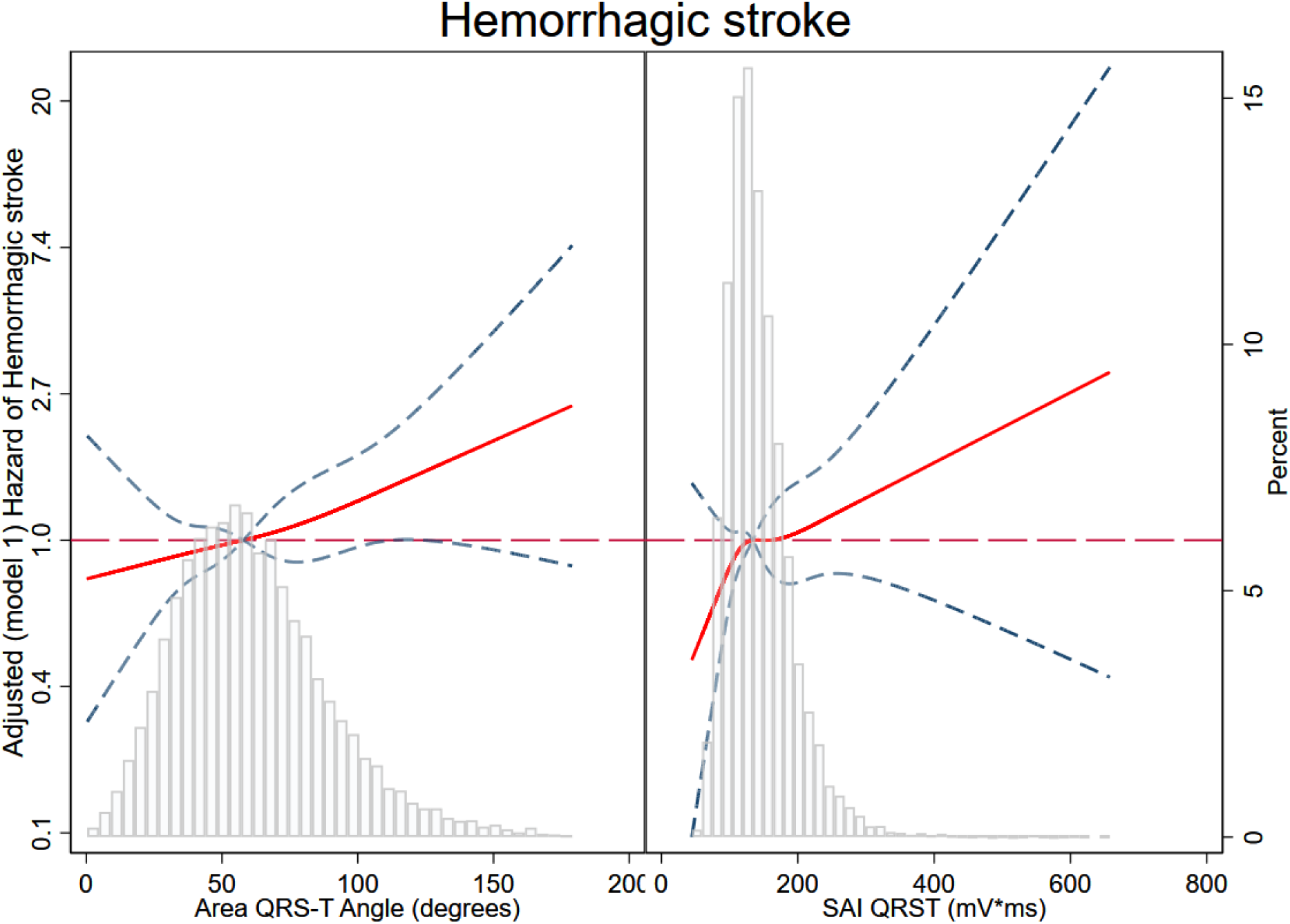
Adjusted (model 1) risk of hemorrhagic stroke associated with area QRS-T angle (A) and SAI QRST (B). See Figure 3 legend for the details.

Notably, the spatial peak QRS-T angle was strongly and independently associated with hemorrhagic stroke across all models (1-5), with little to no attenuation after adjustment for all confounders (Figure 7). Similar to ischemic stroke, SVG magnitude exhibited a time-dependent association with hemorrhagic stroke (Figures 8 and 9). Sensitivity analyses of pure ICH outcome highlighted and strengthened the association of time-updated SVG magnitude with ICH (Tables 4-5).

**Figure 7.**
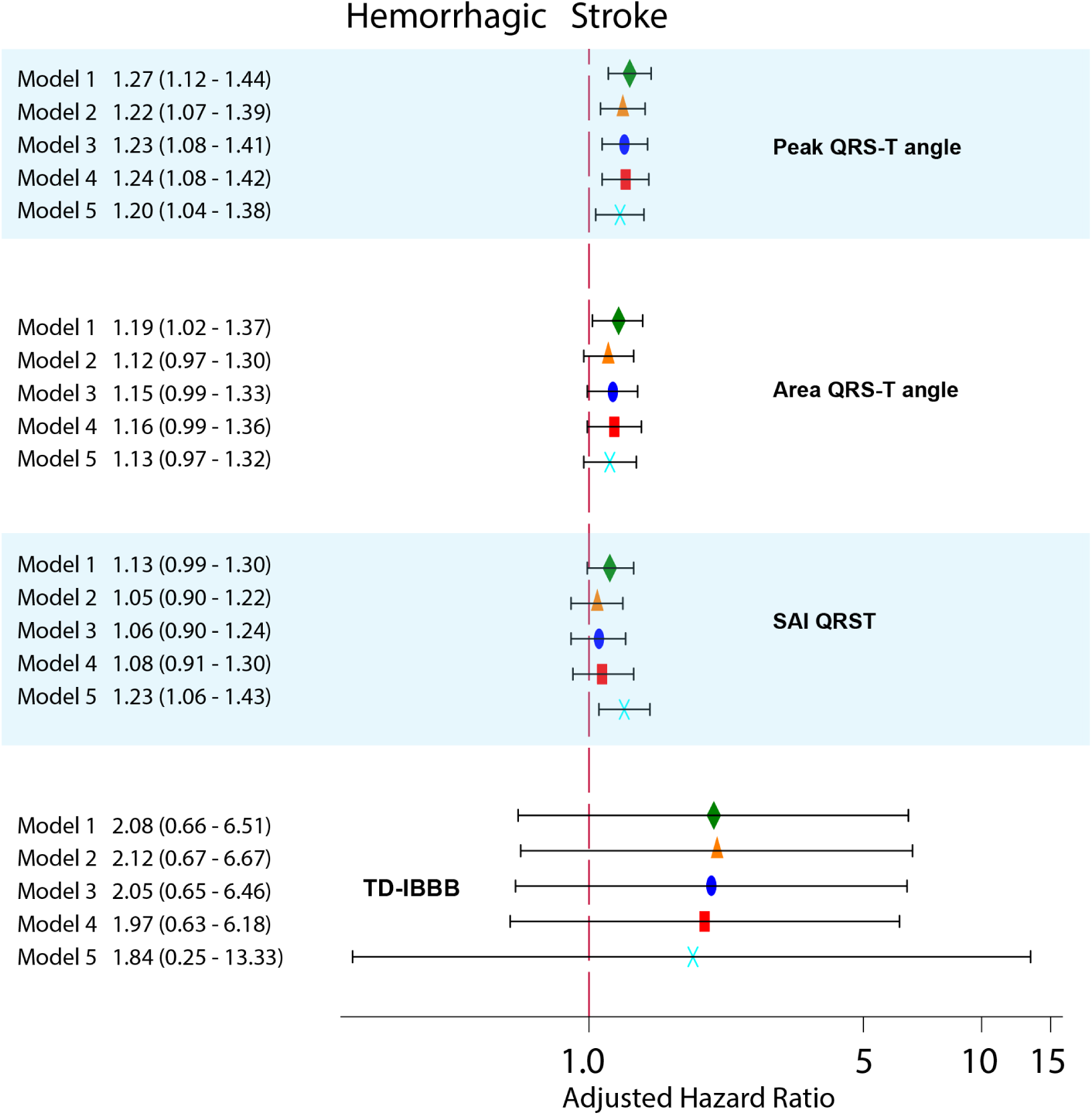
Adjusted Cox proportional hazard ratio (HR) with 95% confidence interval (CI) for the association of peak and area QRS-T angle, SAI QRST, and a TD-IBBB on a 10-second ECG with ICH. See Figure 2 legend for the details. Association of TD-IBBB with pure incident hemorrhagic stroke in Cox models with the sequential adjustment is reported.

**Figure 8.**
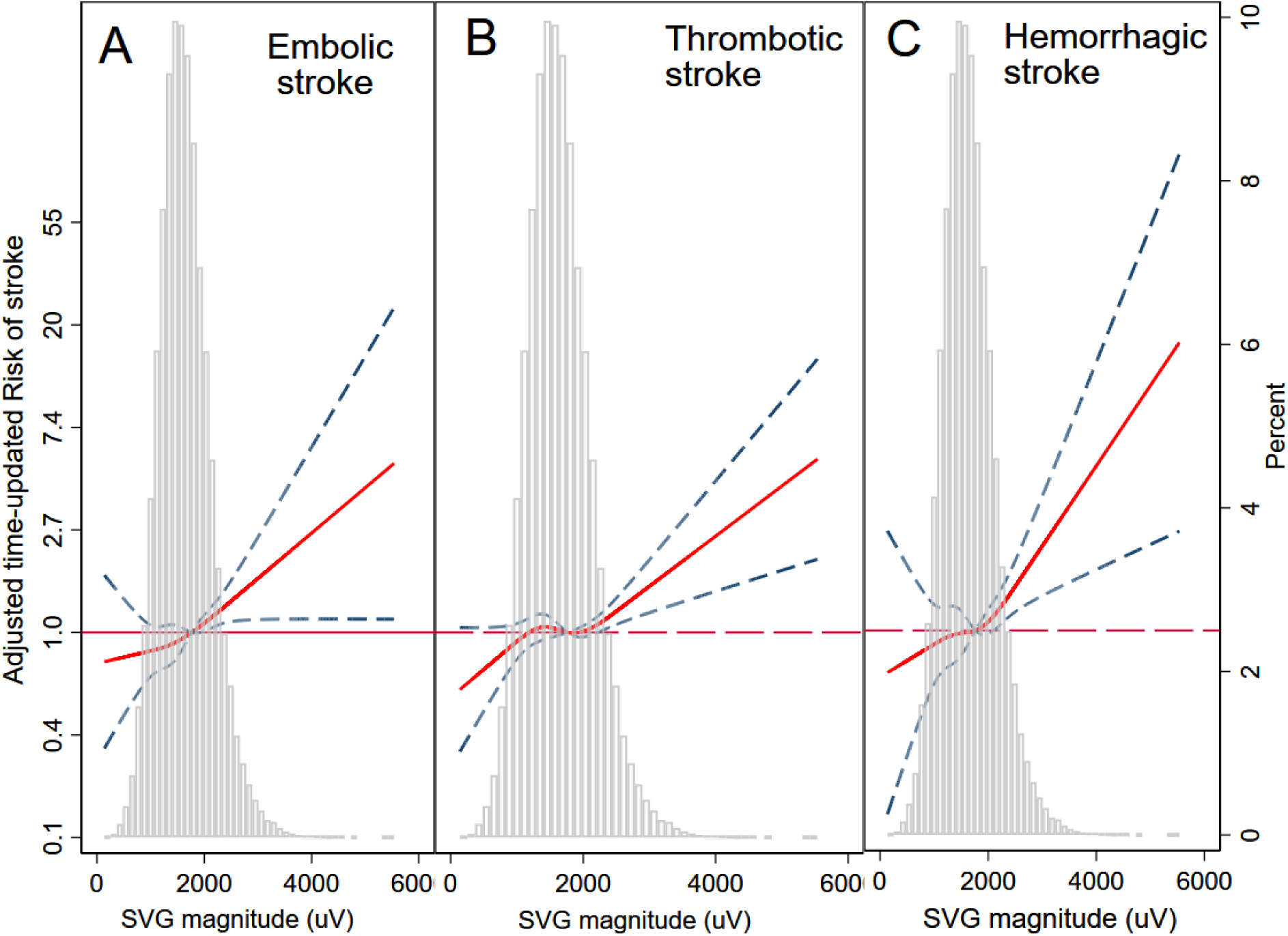
Time-updated adjusted risk of (A) cardioembolic, (B) thrombotic brain infarction, and (C) intracranial hemorrhage associated with SVG magnitude. Time-updated Cox regression models were adjusted for age, sex, race and study center, time-updated heart rate, and the type of median beat. Restricted cubic spline with 95% CI shows a change in the hazard ratio (Y-axis) in response to SVG magnitude change (X-axis). The 50th percentile of SVG magnitude is selected as a reference. Knots of SVG magnitude are at 0.92 – 1.43 – 1.79 – 2.51 mV.

**Figure 9.**
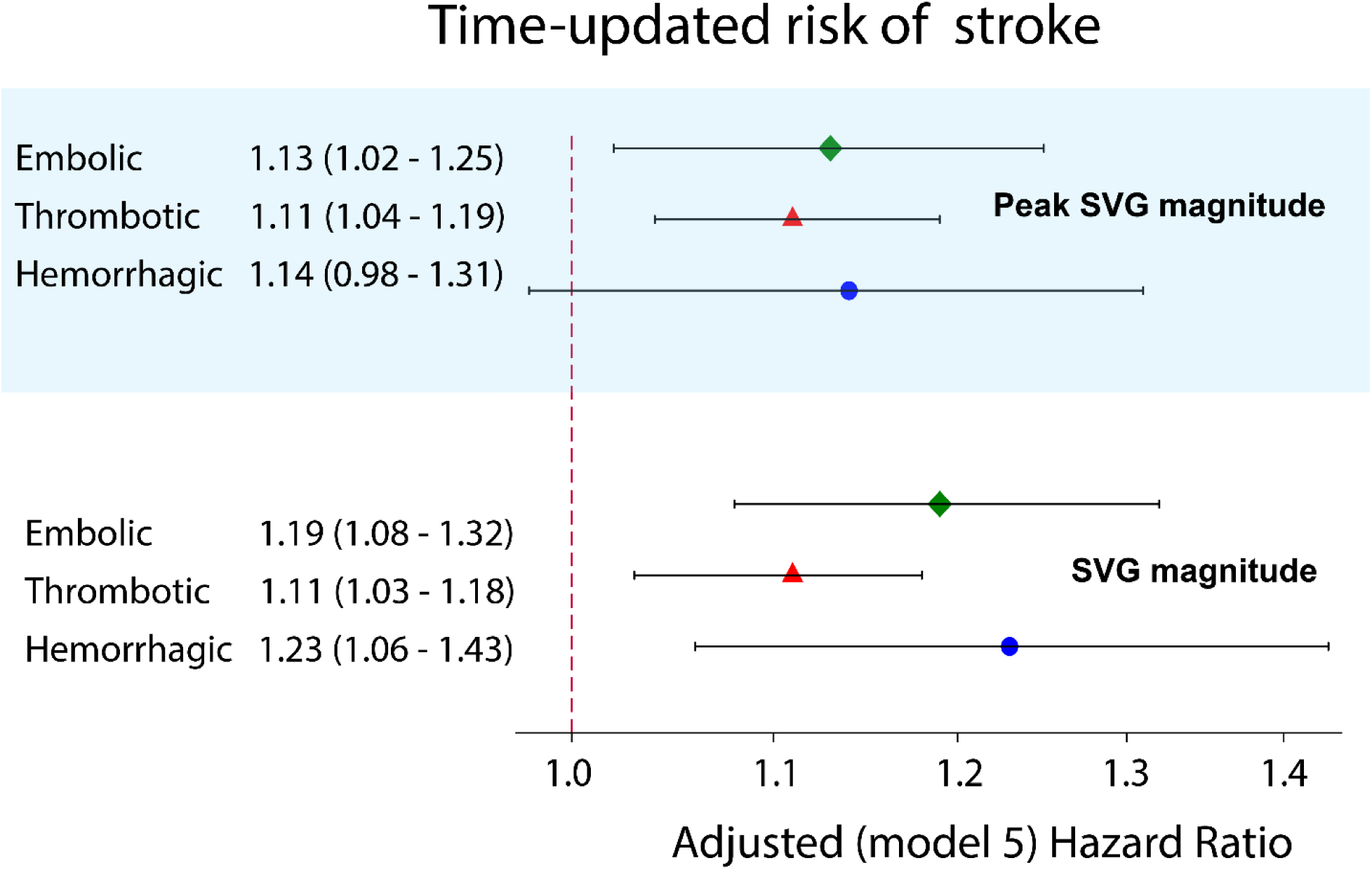
Adjusted Cox proportional hazard ratio (HR) with 95% confidence interval (CI) for the association of SVG magnitude with EBI (green diamond), TBI (red triangle), and ICH (blue circle) in time-updated model 5. See Figure 2 legend for the details.

Similar to thrombotic (but not cardioembolic) stroke, adjustment for atrial substrate did not affect associations of ECG metrics with ICH, suggesting their independence from an atrial substrate. Moreover, similar to cardioembolic (but not thrombotic) stroke, adjustment for traditional ECG measures of ventricular substrate did not affect associations of GEH metrics with ICH, suggesting that they characterize specific features of the ventricular substrate.

As expected, PVCs did not associate with hemorrhagic stroke. There was a nominal association of TD-IBBB with ICH, which was consistent in a time-updated model but did not reach statistical significance (Figure 7 and Tables 4-5). It indicated a nearly two-fold higher risk of ICH in participants with TD-IBBB.

## Discussion

This large, prospective, community-dwelling cohort study of more than 14 thousand adults revealed several novel findings. First, we uncovered CVD-independent ventricular substrate of cardioembolic stroke, likely reflecting the development of cardiac memory in response to idiopathic PVCs. Our results suggest that the search for the sources of cardioembolic stroke should include an assessment of the PVC burden. Our findings open an avenue for randomized clinical trials of the ablation of idiopathic PVCs as a possible prevention strategy for a cardioembolic stroke of uncertain source.

Second, we observed a time-dependent association of a transient ventricular substrate likely reflecting cardiac memory in patients with TBI, suggesting that PVCs can also trigger a thromboembolic stroke from a brain artery. Unlike in cardioembolic stroke, PVCs that are implicated in TBI are more likely associated with CVD substrate. Future studies are needed to determine whether monitoring of cardiac memory (SVG magnitude, SAI QRST, QRS-T angle) and PVC burden in patients at risk for TBI can be useful for determining the timing of carotid endarterectomy and adjustment of the antiplatelet therapy regimen.

Finally, we found an association of CVD-independent ventricular substrate with ICH, likely reflecting the development of cardiac memory in response to TD-IBBB and suggesting an implication of amyloidosis. The TD-IBBB phenomenon and associated cardiac memory in older adults should be studied further. After validation of these study findings in another study, TD-IBBB may be used as a marker of ICH risk, increasing clinical suspicion of cardiac amyloidosis.

Notably, our study suggested that the wide spatial QRS-T angle, and the large SAI QRST and SVG magnitude are non-specific characteristics of the transient substrate of cardiac memory, developing in response to PVCs and TD-IBBB. Further ECG monitoring studies are needed to validate an association of PVC and TD-IBBB burden with the amount of cardiac memory. Monitoring of the amount of cardiac memory on 12-lead ECG can be potentially less burdensome, less expensive, and more efficient than measurement of PVCs and TD-IBBB burden by Holter ECG, or ECG-patch, and should be studied further.

### The ventricular substrate of ischemic stroke

AF is a well-recognized major risk factor, and the central focus for cardioembolic stroke prevention.^44^ The pathophysiological mechanisms of stroke in AF have been studied for decades.^45^ However, a frequently missing temporal link between AF episodes and stroke events remains incompletely understood.^46, 47^ The atrial myopathy hypothesis^48^ was developed to explain the temporal dissociation between AF and stroke, and suggests that the fibrotic atrial substrate, which interacts with the atrial electrical dysfunction^49^, is the principal mediator of thromboembolism. Nevertheless, the risk of stroke is increased in a graded manner in patients with paroxysmal, persistent, and permanent AF.^50, 51^ Moreover, greater AF burden is associated with a higher risk of stroke^52, 53^, highlighting the importance of a hemodynamic disturbance triggering thromboembolic events. Therefore, the current strategy for determining the etiology of cardioembolic stroke is to search for underlying AF.^54^

Our study uncovered the presence of a CVD-independent ventricular substrate associated with cardioembolic stroke (Figure 10), which remained associated with EBI even after rigorous adjustment for CVD and cardiovascular risk factors, AF and atrial substrate^55, 56^, and known ventricular abnormalities (BBB, LVH). PVCs and PVC burden (manifested by the cardiac memory) characterize the CVD-independent ventricular substrate of cardioembolic stroke. Several previous studies reported an association of PVCs on a 2-minute ECG recording with ischemic stroke.^8, 9^ One of these studies, Agarwal et al^8^, had smaller statistical power and did not adjust for AF and atrial substrate, however, their findings were consistent with our own. They observed that PVCs were associated with incident stroke in individuals without hypertension and diabetes, although not in those individuals with hypertension or diabetes. Previous studies^8, 9^, also speculated that PVCs merely reflect the presence and burden of CVD. Nevertheless, our study demonstrated that the association of PVCs with cardioembolic stroke is independent of CVD and cardiovascular risk factors (model 2, Figure 2), as well as traditional CVD-related ventricular substrate (LVH and BBB; model 4, Figure 2). Moreover, we showed that AF and atrial substrate mediate the association of PVCs with cardioembolic stroke (model 3, Figure 2).

**Figure 10.**
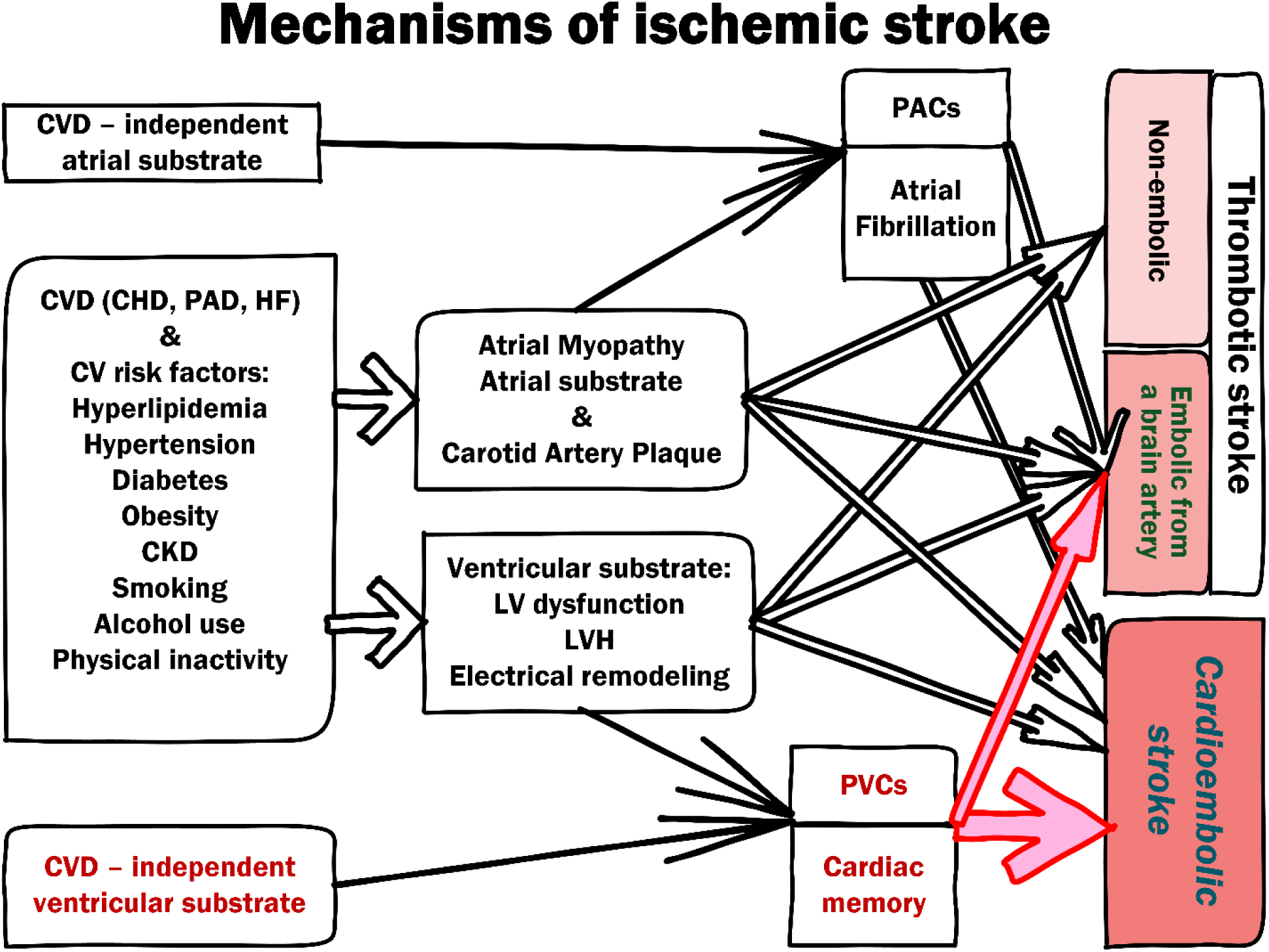
Mechanisms of ischemic stroke. Red color highlights the mechanisms suggested by this study.

It is well-known that PVCs can induce cardiac memory, which is manifested by a wide QRS-T angle^57, 58^, and a large QRST integral.^59^ In this study, the response of the specific ventricular substrate measures (PVCs, spatial QRS-T angle, and SAI QRST) on stepwise adjustment (models 1-4, Figure 2) was similar, supporting the notion that spatial QRS-T angle and SAI QRST reflect cardiac memory. We also observed a gradual, “dose-dependent” association of QRS-T angle and SAI QRST with the hazard of cardioembolic stroke (Figure 3).

Evidently, there are several mechanisms behind abnormal QRS-T angle^60^ and SAI QRST^61^, which explain an attenuation of the association after adjustment for CVD and cardiovascular risk factors. Importantly, spatial QRS-T angle and SAI QRST remained associated with cardioembolic stroke after full adjustment, in time-updated model 5. This raises the possibility that cardiac memory reflects an increasing burden of PVCs before the stoke event. Moreover, we observed a strong, independent, “dose-dependent” (Figure 8) association of time-updated SVG magnitude with cardioembolic stroke. SVG magnitude reflects cardiac memory^15^ as a transient substrate or a trigger.^21^ As expected, the SVG direction did not associate with incident stroke because idiopathic PVCs can originate from different locations.^62^ Therefore, the results of our study suggest that the high burden of PVCs can trigger cardioembolic events. Future studies are needed to prove an association of PVC burden with cardioembolic stroke.

PVC burden can be a “missing” temporal link connecting hemodynamic disturbance due to paroxysmal arrhythmia with acute stroke events in predisposed patients (Figure 10). The ablation of idiopathic PVCs could be an effective and safe^63^ prophylactic measure to reduce the risk of acute stroke. Our findings open an avenue for randomized clinical trials of cardioembolic stroke prevention using the ablation of idiopathic PVCs, especially for the secondary prevention of a cardioembolic stroke of uncertain source.

Consistent with the notion that a subset of TBIs includes embolic strokes originating from a brain artery, we observed a weak association of PVCs, spatial QRS-T angle, SAI QRST, and time-updated SVG magnitude with TBI. As expected and contrary to cardioembolic stroke, the association of a ventricular substrate with TBI was independent of AF and atrial substrate. Still, the association was mediated primarily by CVD and cardiovascular risk factors, including BBB and LVH (Figure 5).

Similar to EBI, a dose-dependent association of spatial QRS-T angle, SAI QRST, and time-updated SVG magnitude with the risk of TBI (Figures 4 and 8) suggested a similar underlying mechanism - cardiac memory, reflecting PVC burden. However, unlike in cardioembolic stroke, an embolic stroke from a brain artery seems to be more likely triggered by CVD-related PVCs. Thus, monitoring of SVG magnitude (i.e., monitoring of cardiac memory due to PVC burden) in patients at risk of TBI can potentially be used to adjust antiplatelet therapy regimen or timing of carotid endarterectomy, which should be investigated further (Figure 10).

### The ventricular substrate in hemorrhagic stroke

In this study, we observed CVD-independent ventricular substrate of ICH that similar to the ventricular substrates of EBI and TBI characterizes cardiac memory (wider QRS-T angle, larger SAI QRST and transient increase in SVG magnitude). However, unlike in ischemic stroke, in hemorrhagic stroke, cardiac memory is developed in response to TD-IBBB^64^, suggesting the implication of amyloid (Figure 11).

**Figure 11.**
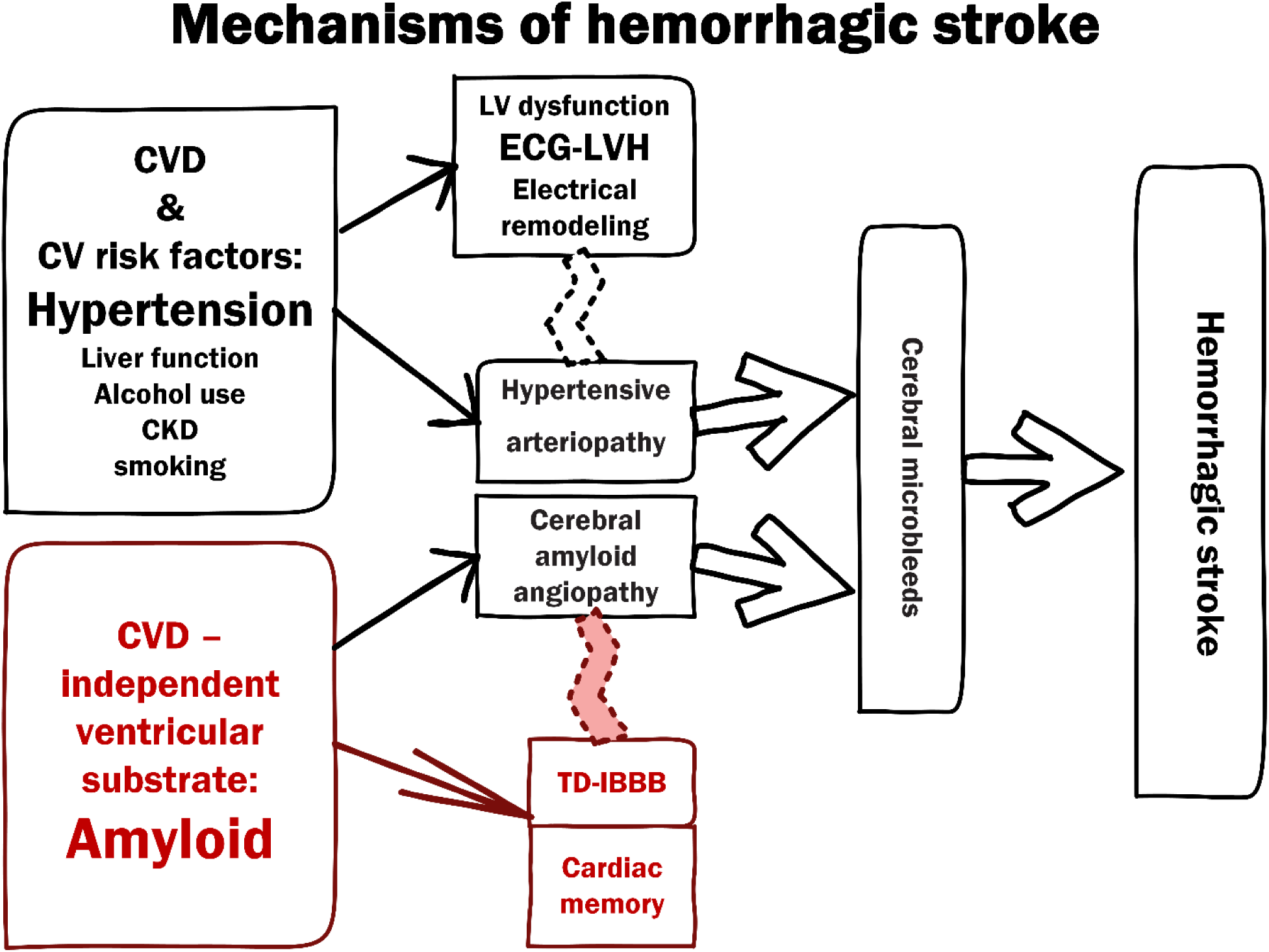
Mechanisms of hemorrhagic stroke. Red color highlights the mechanisms proposed by this study.

Cerebral microbleeds are the strongest risk factors of hemorrhagic stroke.^65^ Cerebral amyloid angiopathy is the second most common cause of hemorrhagic stroke, following hypertension (hypertensive arteriopathy).^66^ The previous study demonstrated an association of ECG-LVH with ICH.^36^ We adjusted our analyses by ECG-LVH, history of hypertension, antihypertensive treatment, and levels of systolic and diastolic blood pressure, to account for hypertensive arteriopathy as the leading cause of ICH.

The amyloid burden is independently associated with cerebral microbleeds.^67^ Amyloidosis is a systemic disease. Cardiac amyloidosis is significantly underdiagnosed, partly because of a lack of suspicion and a widespread belief that it is rare.^68^ Nevertheless, wild-type transthyretin (ATTRwt) cardiac amyloidosis is common in older adults^69^ and HF patients with preserved ejection fraction.^70^ Recently, novel diagnostic approaches^71^ and therapies^72^ emerged, which can improve clinical outcomes but also demonstrate complications of longer disease durations^73^.

Previously described ECG manifestations of cardiac amyloidosis are highly variable^74^ and include ECG-LVH, low voltage, and poor R wave progression.^70^ Notably, one consistent characteristic feature of all types of cardiac amyloid (light chain, mutant ATTR, and ATTRwt) includes cardiac conduction abnormalities.^75^ In this study, individuals with TD-IBBB had a two-fold higher risk of ICH, but the association was not statistically significant and should be validated in another independent study.

Up to 25% of patients with cerebral amyloid angiopathy are also diagnosed with AF^76^, which poses a difficult dilemma and discussion about the risks and benefits of anticoagulation for stroke prevention.^77^ In our study, 19 patients experienced both hemorrhagic and ischemic stroke. Our results hold a promise that in the future propensities towards ischemic and hemorrhagic stroke can be dissected based on the presence of TD-IBBB or PVCs, and monitoring the extent of cardiac memory.

### Strengths and Limitations

The strengths of our study included a large prospective cohort with long-term follow-up and well-adjudicated stroke events, providing sufficient statistical power for rigorous adjustment. However, the limitations of the study should be taken into consideration. While the statistical power for ischemic stroke was adequate, there were fewer ICH events, which challenged a fair comparison of models for ischemic and hemorrhagic stroke events. To overcome that limitation, we conducted sensitivity analysis and utilized sequential rather than an incremental adjustment in Cox regression models with ICH outcome.

In 2017, the American College of Cardiology/American Heart Association Guideline lowered the threshold for the definition of hypertension in adults (systolic blood pressure ≥130 or a diastolic blood pressure ≥80 mmHg)^78^. To maintain consistency with all previous research in ARIC, we kept the historical definition of hypertension. However, we also adjusted for actual blood pressure values, considering the paramount importance of hypertension as a stroke risk factor.

Assessment of paroxysmal arrhythmic events (AF, PACs, PVCs, TD-IBBB) on a 10-second ECG cannot accurately represent the burden of arrhythmia. To minimize this limitation, we assumed that AF diagnosed at any time during follow-up, as well as PACs, PVCs, and TD-IBBB, detected at any visit have been present since baseline. Moreover, we also assessed PACs, PVCs, and TD-IBBB in time-updated models, updating their presence on the exact day of ECG recording.

## Data Availability

The ARIC Study data are available through the National Heart, Lung, and Blood Institute's Biological Specimen and Data Repository Information Coordinating Center (BioLINCC), the National Center of Biotechnology Information's database of Genotypes and Phenotypes (dbGaP), and via ARIC Coordinating Center at the University of North Carolina-Chapel Hill.

## Acknowledgments

The authors thank the staff and participants of the ARIC study for their important contributions.

---

**Supplemental Figure 1.**
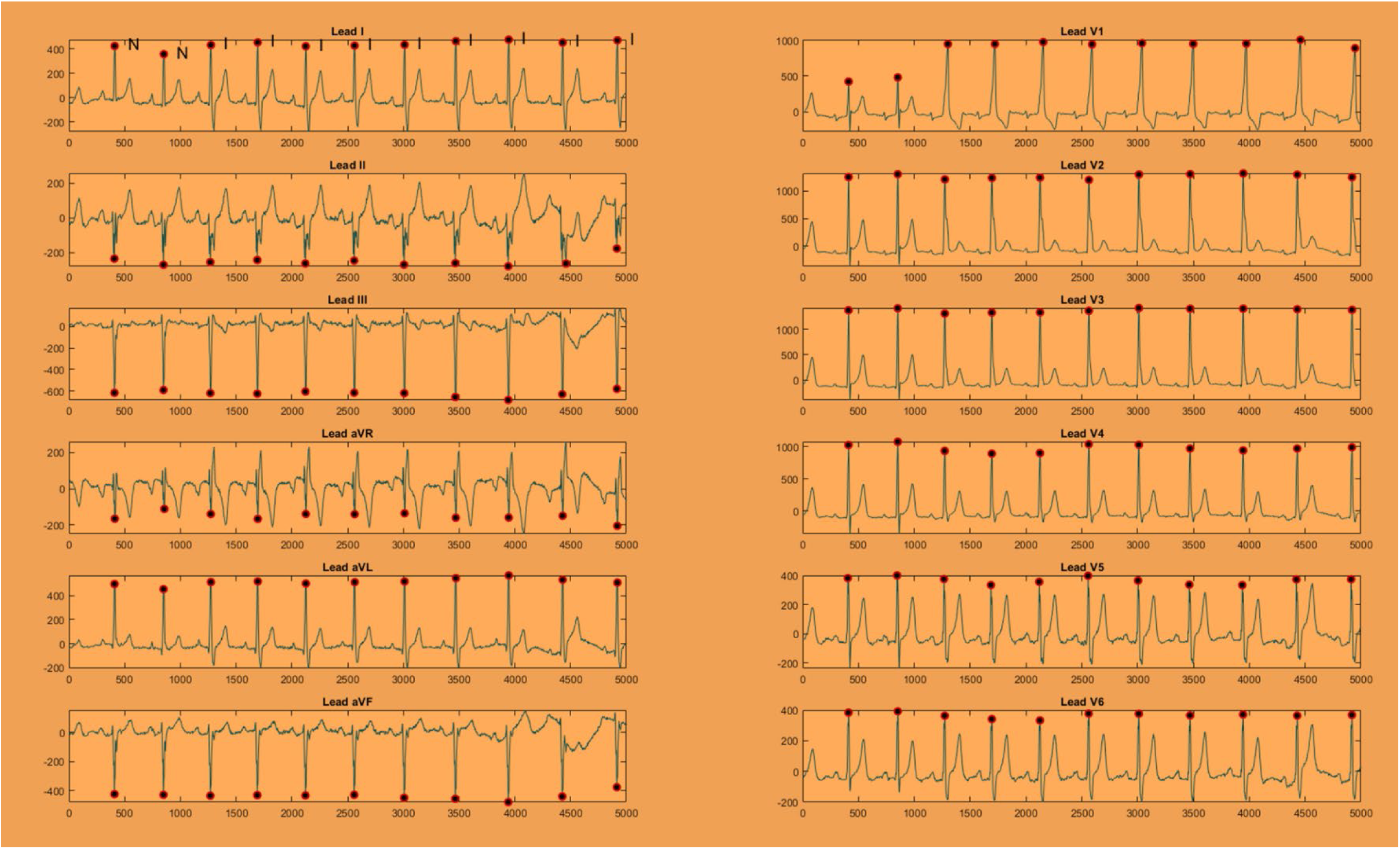
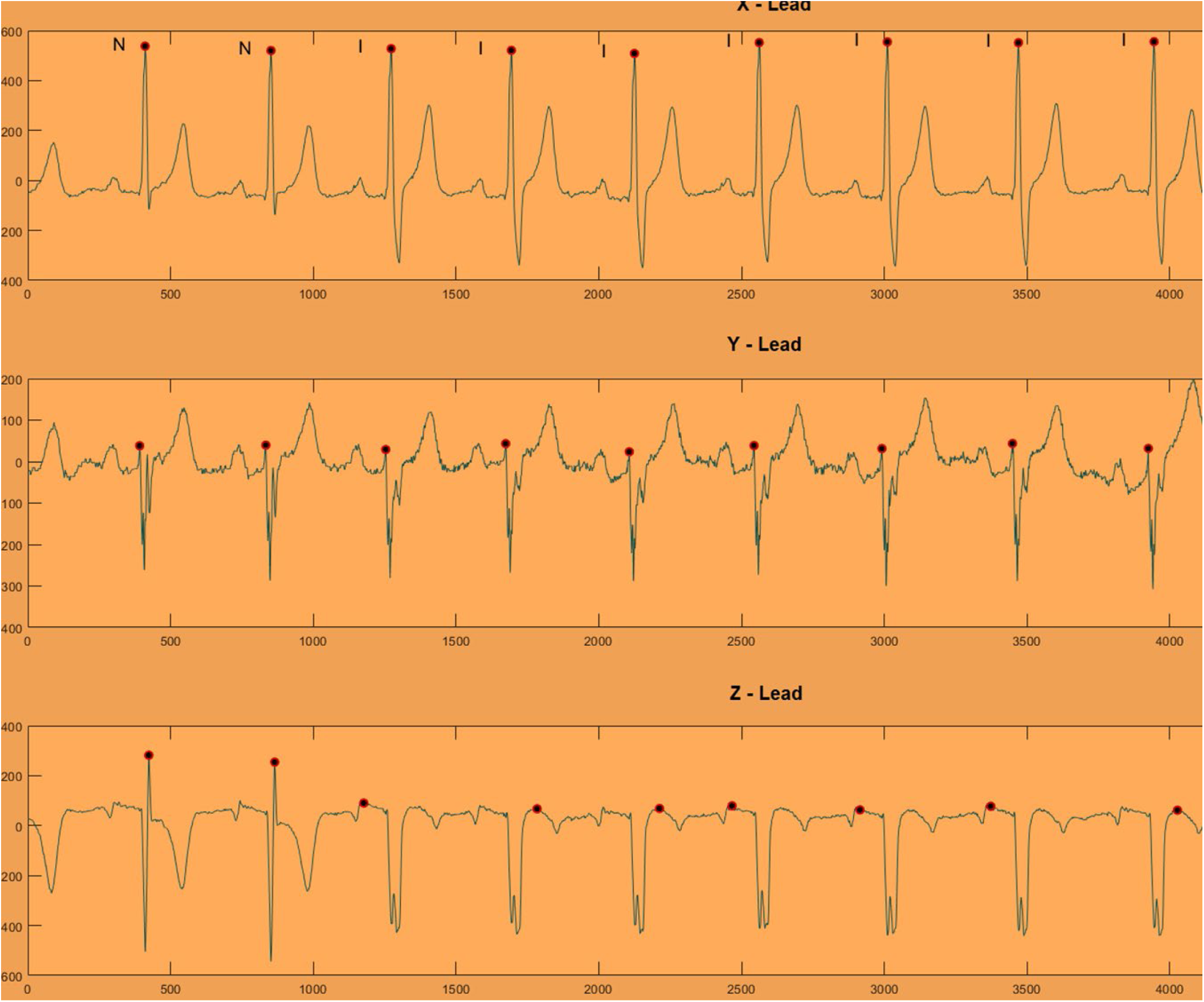
A. Representative example of tachycardia-dependent intermittent right bundle branch block on a 12-lead ECG (A) and orthogonal XYZ ECG (B). Normal sinus beats are labeled N. Sinus beats with intermittent bundle branch block beats are labeled I.

**Supplemental Figure 2.**
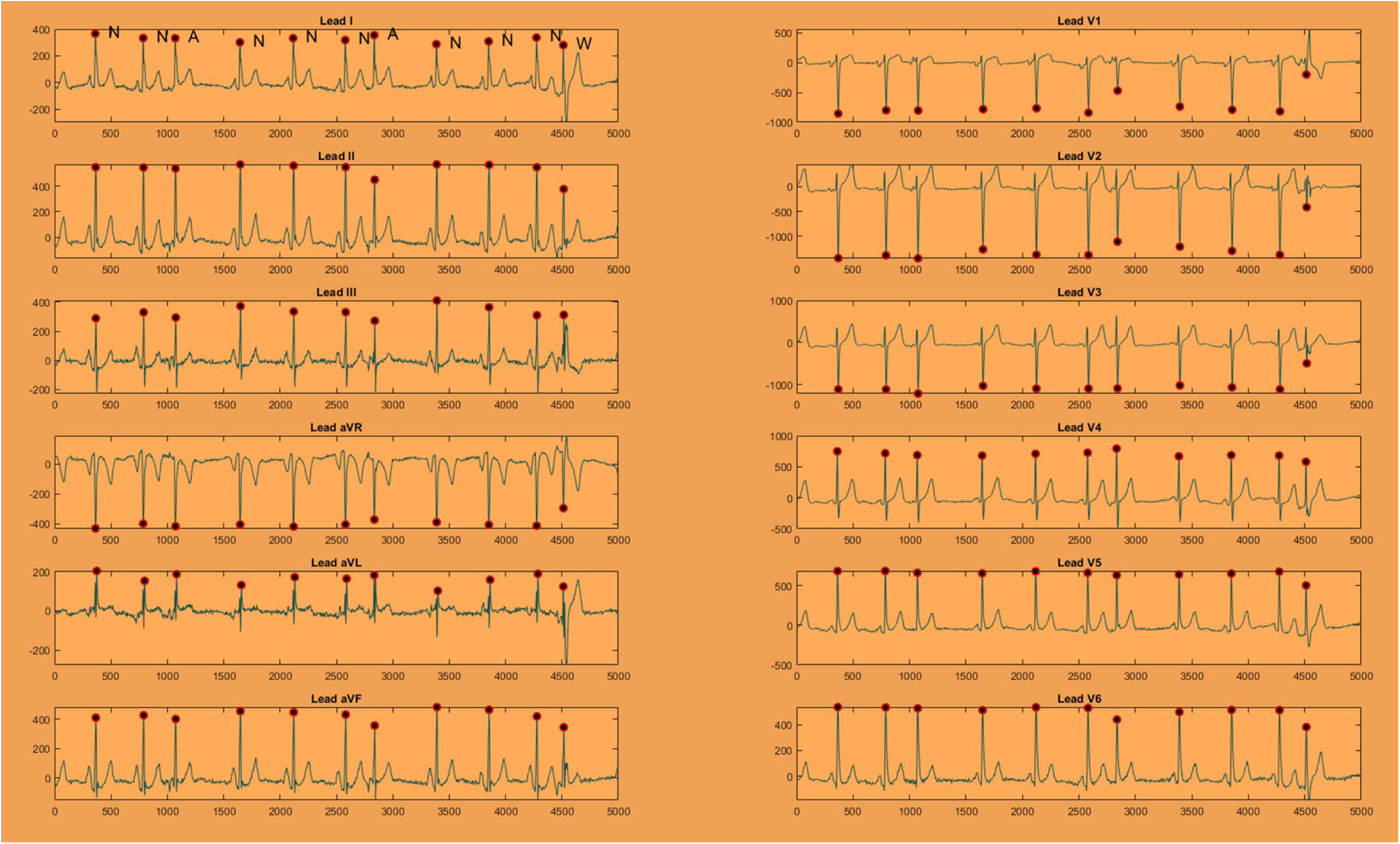
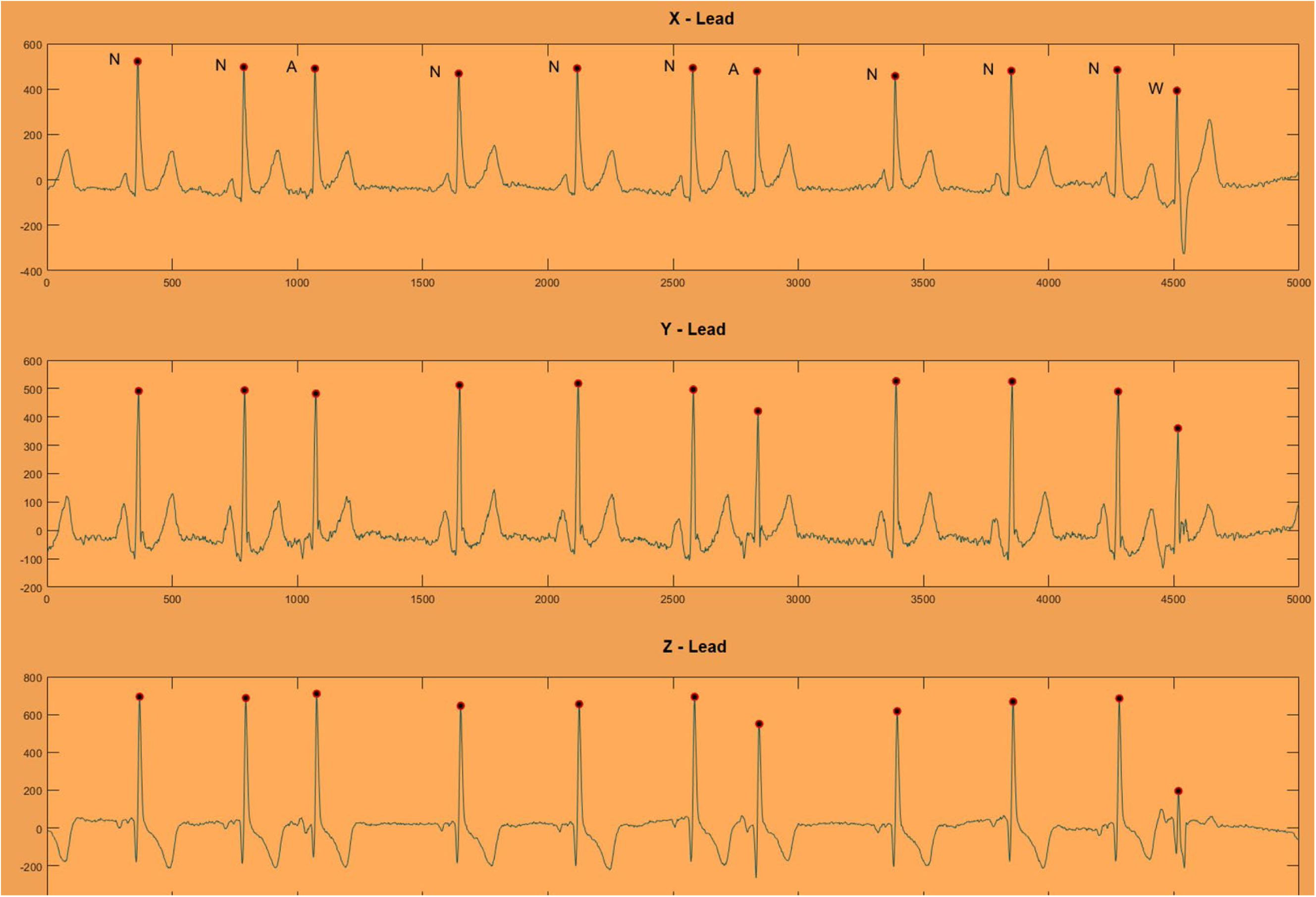
A representative example of an aberrant premature atrial complex on a 12-lead ECG (A) and orthogonal XYZ ECG (B). Normal sinus beats are labeled N. Premature atrial complexes with normal ventricular conduction are labeled A. Premature atrial complex with aberrant ventricular conduction (right bundle branch block) is labeled W.

